# Interrater agreement of annotations of epileptiform discharges and its impact on deep learning – A pilot study

**DOI:** 10.1101/2024.04.10.24305602

**Authors:** Mats Svantesson, Anders Eklund, Magnus Thordstein

## Abstract

1.

**Background:** Expert interrater agreement for epileptiform discharges can be moderate. This reasonably will affect the performance when developing classifiers based on annotations performed by experts. In addition, evaluation of classifier performance will be difficult since the ground truth will have a variability. In this pilot study, these aspects were investigated to evaluate the feasibility of conducting a larger study on the subject.

**Methods:** A multi-channel EEG of 78 minutes duration with abundant periodic discharges was independently annotated for epileptiform discharges by two experts. Based on this, several deep learning classifiers were developed which in turn produced new annotations. The agreements of all annotations were evaluated by pairwise comparisons using Cohen’s kappa and Gwet’s AC1. A cluster analysis was performed on all periodic discharges using a newly developed version of parametric t-SNE to assess the similarity between annotations.

**Results:** The Cohen’s kappa values were 0.53 for the experts, 0.52–0.65 when comparing the experts to the classifiers, and 0.67–0.82 for the classifiers. The Gwet’s AC1 values were 0.92 for the experts, 0.92–0.94 when comparing the experts to the classifiers, and 0.94–0.96 for the classifiers. Although there were differences between all annotations regarding which discharges that had been selected as epileptiform, the selected discharges were mostly similar according to the cluster analysis. Almost all identified epileptiform discharges by the classifiers were also periodic discharges.

**Conclusions:** There was a discrepancy between agreement scores produced by Cohen’s kappa and Gwet’s AC1. This was probably due to the skewed prevalence of epileptiform discharges, which only constitutes a small part of the whole EEG. Gwet’s AC1 is often considered the better option and the results would then indicate an almost perfect agreement. However, this conclusion is questioned when considering the number of differently classified discharges. The difference in annotation between experts affected the learning of the classifiers, but the cluster analysis indicates that all annotations were relatively similar. The difference between experts and classifiers is speculated to be partly due to intrarater variability of the experts, and partly due to underperformance of the classifiers. For a larger study, in addition to using more experts, intrarater agreement should be assessed, the classifiers can be further optimized, and the cluster method hopefully be further improved.

## 2. Introduction

Automating tasks that are time-consuming and depend on expert knowledge is important to increase availability. This is certainly the case in EEG analysis and deep learning has emerged as a promising way to accomplish this (Craik et al., 2019; Roy et al., 2019). To assess the utility and risks of the technology, it is important to evaluate it, as well as to study different aspects of the conditions for developing deep learning, to optimize the performance. Most deep learning applications for detecting epileptiform discharges (EDs) are developed using supervised learning (Nhu et al., 2022) where deep classifiers are trained to align to human expert assessments. At present time, there is no generally accepted objective way of defining or quantitatively measure EDs and the gold standard is rating by experts. However, experts do not always agree and in this work, interrater agreement for EDs and how it affects training of deep classifiers was investigated.

In cases where there is no way to know which expert is right, one can at least evaluate if how the experts are assessing is reproducible or how much they agree. The most frequently used quantitative measures for the agreement between experts in the setting of rating EDs, as found in the review below, are percent agreement, Cohen’s kappa (Cohen, 1960), Fleiss’ kappa (Fleiss, 1971), and Gwet’s AC1 (Gwet, 2008). The percent agreement is simply the percentage of the data for which the experts agree. Cohen’s kappa, Fleiss’ kappa, and Gwet’s AC1 produce relative scores where the percent agreement is adjusted for the agreement by chance. Cohen’s kappa uses the marginal distribution to estimate the chance agreement (Cohen, 1960) and Fleiss’ kappa is an extension of this to multiple raters. These two kappa scores have been criticized for producing unexpectedly low scores in some cases of high percent agreement and higher scores when the marginal distribution is asymmetrical compared to the symmetrical case (Gwet, 2014). To address the problems with Cohen’s kappa, Gwet (2008) suggests an agreement coefficient, now referred to as Gwet’s AC1, which is less sensitive to variation in the marginal distributions. To interpret the kappa scores, Landis and Koch (1977) suggest: 0–0.20, slight agreement; 0.2–0.40, fair agreement; 0.4–0.60, moderate agreement; 0.61–0.80, substantial agreement; and 0.81–1.00, almost perfect agreement.

Studies assessing interrater agreement for EEGs can be divided into the rating of individual EDs and rating the presence of EDs in whole EEGs. In the former case, the median agreement score is 0.56 (min: 0.30; max: 0.81) (Bagheri et al., 2017; Beuchet et al., 2017; Dümpelmann and Elger, 1999; Halford et al., 2017; Halford et al., 2018; Jing et al, 2019; Wilson et al., 1996). In the latter case, the median score is 0.67 (0.16, 0.83) (Abend et al., 2011; Abend et al., 2017; Azuma et al., 2003; Black et al., 2000; Gaspard et al. 2014; Hussein et al., 2017; Mani et al., 2012; Nguyen et al., 2010; Piconelli et al., 2005; Reus et al., 2022; Stroink et al., 2006; Zhuo et al., 2019). The median agreement is thus higher for whole EEGs, but the number of studies is relatively low, they use different types of agreement scores, the number of raters vary from 2 to 49, which makes the difference difficult to assess. A few studies compare the two conditions, and the scores are higher when assessing whole EEGs (Black et al., 2000; Jing et. al., 2019). In summary, the agreement for EDs has a large variation and there are methodological differences that make comparisons of different studies difficult, but overall, the agreement is moderate for individual EDs and possibly substantial for the presence of EDs in whole EEGs.

Expert annotations are used as labels in classification tasks in the setting of supervised learning in deep learning. From what has been presented above, there will thus be a label noise due to disagreement between experts for EDs. It is known that label noise has a negative impact on training deep classifiers in medical imaging (Karimi et al., 2020; Abdala and Fine, 2023). The subject has to the best of our knowledge not been thoroughly investigated in clinical neurophysiology, but it is reasonably a general problem. Label noise will also make evaluation harder since it is not known if the labels are correct—an error in the prediction accuracy may be either due to an error made by the expert or the classifier. When reviewing studies where deep learning is applied to ED classification, the most used performance measures are accuracy (ACC) and area under the curve (AUC). Including studies using either or both of these measures, twenty-two relevant studies were found spanning the years 2016–2023. The median ACC is 0.90 (min: 0.71; max: 1.00) and the median AUC is 0.94 (0.76, 1.00) (Abou Jaoude et al., 2020;Antonaides et al., 2017; Cheng et al., 2022; Chung et al., 2023; Faghihpirayesh et al., 2021; Fürbass et al., 2020; Geng et al., 2021; Jeon et al., 2022; Jiang et al., 2023; Johansen et al., 2016; Kural et al., 2022; McDougall et al., 2023; Medvedev et al., 2019; Nejedly et al., 2023; Nhu et al., 2023; Thangavel et al., 2021; Thomas et al., 2020; Thomas et al., 2021; Tjepkema-Cloostermans et al., 2018; Wang et al., 2023; Wei et al., 2021; Zhang et al., 2023). In these studies, there is a large variation in data sizes, in the methods to calculate ACC (e.g., some used balanced ACC) and AUC (e.g., ROC vs. precision and recall), making comparisons difficult.

The commercial system Persyst 13 has been evaluated in several studies, and its performance in detecting EDs is often characterized as ‘noninferior’ compared to experts (Halford et al., 2018; Joshi et al., 2018; Kural et al., 2022; Reus et al., 2020; Reus et al., 2021; Reus et al., 2022), but it is unclear to what extent it is based on deep learning. Searching the literature, no other studies were found where both interrater agreement for EDs between experts and between experts and deep learning classifiers are assessed. Wang et al. (2023) report an AUC of 0.99 for their classifier and a Cohen’s kappa of 0.91 when comparing it to three experts. The classifier developed by Thomas et al. (2020) achieve an AUC of 0.84 and percent agreement of 81 with one expert. In addition, in the study by Abou Jaoude et al. (2020), they report that 36 percent of false positives made by their classifier are retrospectively assessed as EDs missed by the annotating expert.

In this pilot study, two experts annotated EDs, and different classifiers were created based on their annotations. The agreements between the experts’ and classifiers’ annotations were assessed and the results were also visualized using a newly developed clustering technique (Svantesson et al, 2023). The main aim was to evaluate the feasibility of performing a larger study.

## 3. Materials and methods

### 3.1. EEG data

One EEG from the Temple University Hospital EEG Corpus database (Obeid and Picone, 2016) was used for this study. The EEG had been extracted previously in the context of another study (Svantesson et al., 2021). The duration was approximately 78 minutes sampled at 250 Hz. Electrodes were placed according to the international 10–20 system. Twenty-one electrodes were used (Fp1, Fp2, Fz, F3, F4, F7, F8, Cz, C3, C4, Pz, P3, P4, T3, T4, T7, T8, O1, O2, A1, and A2).

The EEG represented a focal status epilepticus with continuous periodic discharges (PDs) with varying morphology, of which a subset was epileptiform (Fig. 1). It was selected for the study because the exam was somewhat longer than a standard exam, it contained relatively large amounts of candidate EDs which had a variation in morphology but relatively similar electric fields. The expectation was that this would provide enough data for training and analysis, while still being a reasonable task for the experts to perform.

**Fig. 1.**
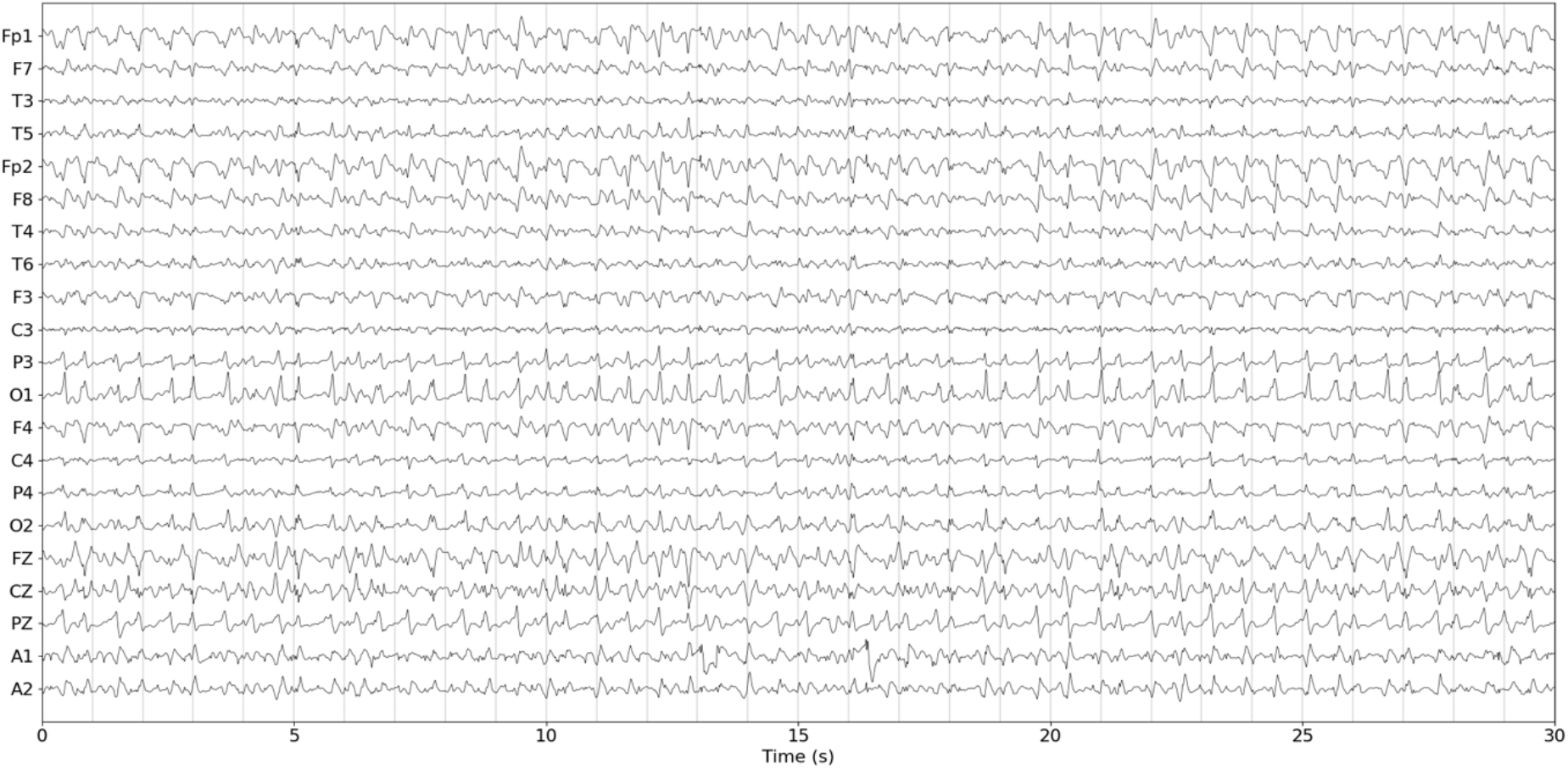
Thirty seconds excerpt from the EEG in common average reference.

#### 3.1.1. Preprocessing

For the visualization, the EEG was bandpass filtered between 0.3 and 40 Hz. For deep learning classifiers and cluster encoders, the EEG was bandpass filtered between 1 and 40 Hz, re-referenced to the common average, and normalized by dividing by the 99^th^ percentile of the absolute amplitude. All filters were implemented in Python as 5^th^ order Butterworth filters using scipy.signal (Virtanen et al., 2020) and zero-phase filtering.

#### 3.1.2. EEG annotation

Two senior specialists in clinical neurophysiology (author M.T. and another member of the research group, non-author; both with more than 20 years of experience) independently annotated the EEG for EDs. They were instructed to only annotate potentials with a strict epileptiform morphology, where each potential should be assessed independent of other potentials. No further instructions or definitions were provided of what constitutes EDs. From their annotations, two additional annotations were created by taking the union and the intersection of the two original annotations.

The EEG was also annotated for PDs (author M.S., specialist in clinical neurophysiology). It was assumed that the same biological process generated all the PDs, i.e., discharges of epileptiform morphology were not generated by another process. The intention was thus to let the epileptiform discharges be a subset of the PDs and to guarantee this, the union of all annotations were taken as the final annotation of PDs.

#### 3.1.3. Visualization and manual annotation

A GUI implemented in Python was created for visualizing and annotating the EEG. The visualization was calibrated to display amplitude as 100 μV per cm and time as one second per three cm. Fifteen seconds of EEG was displayed at a time. It was possible to navigate forward or backward in steps of 1 or 15 seconds, and to jump directly to the start or the end. Navigation was possible by using the mouse and clicking on buttons in the GUI, or by using designated buttons on the keyboard. The EEG could be visualized in a common average reference or a longitudinal bipolar montage.

EDs were annotated by using the mouse cursor to place markings. The experts were instructed to position the markers at the peak of the maximum of each discharge. Each marker only represented the time of the discharge, not which EEG-lead the maximum was located in. Each annotation was saved as an array of zeros and ones, where ones signified EDs. The length of the arrays was equal to the length of the EEG (78 minutes sampled at 250 Hz). For each marking the 29 preceding and 30 succeeding positions in the array were set to one. Each EDs would then have a length of 60 sample points or 240 ms.

### 3.2. Deep learning

All deep learning was developed in Python (version 3.7.13) using the API Keras (version 2.8.0) and the programming module TensorFlow (version 2.8.0). Three different computers were used, equipped with 64–128 GB of RAM, and Nvidia graphics cards (Quadro P5000, Quadro P5000 RTX, or two Titan RTX).

The Adam optimizer was used in all training (Kingma and Ba, 2015) with parameters β1 = 0.5 and β2 = 0.9. A learning rate of 1e-5 was used for the classifiers and 1e-4 for the cluster encoder. Batch sizes of 500 were used in all instances. The binary cross-entropy was used as loss function for the classifiers. To reduce differences due to random variations, the classifiers were initialized using the same values for the weights.

In what follows, channel refers to EEG-channel.

#### 3.2.1. Classifiers

All classifiers used the same architecture (Fig. 2) and hyperparameters. The input size to the classifiers were 21×60 (channels x time). Each classifier started with five convolutional layers; each layer was followed by a scaled-exponential rectifying unit (SELU) (Klambauer et al., 2017), and a max pooling layer which downsampled by a factor of two. Filter kernel sizes were three and the number of filters per layer were 32, 64, 128, 256, and 512. Channels were analyzed separately. Thereafter, four fully connected layers followed, the three first having 512 nodes. SELU activations was used for all layers except the last, which had one node and a sigmoid activation to complete the classifier. The total number of trainable parameters was 6,640,769.

**Fig. 2.**
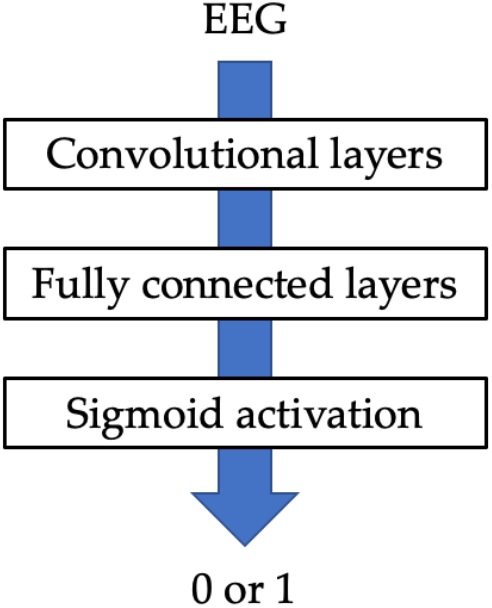
Classifier architecture. The classifier starts with a series of five convolutional layers. Each convolutional layer is followed by a SELU activation and a max pooling layer using strides of two. This is followed by three fully connected layers with SELU activations in between. The classifier ends in a fully connected layer with one node followed by a sigmoid activation. The value of the output is rounded off relative a threshold to produce a value of 0 or 1, where 0 signifies no ED and 1 signifies ED.

Due to the small data size, a five-fold cross-validation strategy was used in the evaluation. The EEG was partitioned into five segments, training was performed on four segments with evaluation on the remaining segment, and this was repeated for all five combinations so that the whole EEG was evaluated.

Using a validation set in every instance to determine when to end training proved difficult due to large variation in validation values from iteration to iteration. Instead, repeated training was performed using the annotation of expert 1, withholding 15 percent of the intended training data as validation data, and then averaging the validation loss and accuracy. From these tests, it was determined that 3,000 iterations (corresponding to roughly 12 minutes of training for the used computer) was adequate to maximize the accuracy of the validation data without exaggerating the overfitting. Comparisons were also made between using different distributions of the three categories: non-ED/non-periodic, non-ED/periodic, and ED data. The large variation in validation output made comparisons difficult, but there did not seem to be a distribution that would produce a high accuracy without producing many false positives and minimizing false positives or both false positives and negatives yielded low number of true positives. In the end, it was decided towards aiming to detect an equal amount of EDs as the respective annotation since the aim of the study was to compare the annotations. A distribution of 0.15, 0.75, and 0.1 (non-ED/non-periodic, non-ED/periodic, and ED) was therefore used for the final results. It was not tested to use different weights for the categories in the loss function.

To produce as much variation as possible in the training data, batches of examples were generated by random selection from the EEG. Each example was selected by first randomly selecting one of the three categories defined above according to a given distribution, and then generating a random position in the EEG that belonged to the selected category. An example was defined as ED if > 75 percent overlapped with an ED marking, as periodic non-ED if not ED and > 75 percent overlapped with a periodic marker, and as non-periodic/non-ED otherwise. The input size of 60 samples was chosen so that the EDs would fit inside while still allowing some variation where the EDs were located (cf., Fig. 18) and without risk of overlap between EDs (or PDs).

Since EEG is continuous data where EDs can occur at any time, the training and test data was for each classifier scanned in steps of 15 samples, i.e., overlapping. An array of ones and zeros was created where each segment classified as ED was marked as ones (60 sample steps), and the remaining array positions were marked as zeros. The scanning process made some markings longer than 60 sample steps (75 or 90 steps). The evaluation time for each fold was 45–50 minutes.

#### 3.2.2. Cluster encoder

Cluster analysis was performed on the PDs, to better understand the results. A further development of a deep learning-based cluster encoding published previously (Svantesson et al., 2022) was used. The clustering is based on the principle of t-distributed stochastic neighbor embeddings (t-SNE) where high-dimensional data can be reduced into two dimensions to produce visualizations (van det Maaten and Hinton, 2008). The improved version used here, produced more distinctly separate clusters and in addition produced identifiers for each cluster. Details on the cluster encoder developed for this work is presented in Appendix 9.3.

### 3.3. Statistics

In the evaluation, the data were analyzed as consecutive examples of 240 ms duration. The whole EEG then amounted to 19,408 examples. When comparisons were made between different annotations, overlapping markers corresponding to the same potentials had to be aligned since the exact position would vary between both experts and classifiers. The result of this procedure was visually inspected by plotting the original and aligned markers across the EEG to ensure accurate results.

In previous studies, presented in the introduction, there is a large variation in used metrics to assess the accuracy of classifiers. In this pilot study a battery of metrics was included. In addition to ACC and AUC based on the receiver operating characteristic (R-AUC), balanced accuracy (B-ACC), AUC based on precision-recall (PR-AUC), F0.5, and Matthew’s correlation coefficient (MCC) were used. The library sklearn.metrics (Pedregosa et al., 2011) was used to calculate these metrics. For R-AUC, average was set to “weighted”. For F0.5, average was set to “binary”. Sensitivity, specificity, precision, and negative predictive value (NPV) was also calculated.

Interrater agreement was assessed by pairwise comparisons using both Cohen’s kappa and Gwet’s AC1. Cohen’s kappa was computed using sklearn.metrics.cohens_kappa_score. Gwet’s AC1 was computed using irrCAC (https://pypi.org/project/irrcac/; accessed 2023-10-15).

## 4. Results

### 4.1. Annotations

Expert 1 identified 1,709 EDs, expert 2 identified 1,430 EDs, the union of expert 1 and 2 resulted in 2,253 identified EDs, and the intersection of expert 1 and 2 resulted in 886 identified EDs. There were 8,107 identified PDs. Examples of marked potentials are presented in the Appendix, section 9.1. All experts used the average referenced montage, and the annotation time was about two hours.

### 4.2. Classifiers

For the classifiers to identify roughly the same number of EDs as their respective expert annotation they were trained on, the thresholds for identification based on the sigmoid output were adjusted to 0.48, 0.45, 0.35, and 0.55.

The performances as assessed by the metrics were moderate in most instances (Tab. 1): B-ACC 0.76–0.82, R-AUC 0.52–0.72, PR-AUC 0.51–0.65, F0.5 0.53–0.75, MCC 0.51–0.72, sensitivity 0.54–0.75, and precision 0.55–0.75. Most of the EEG was non-ED, and this was reflected in higher scores for ACC 0.94–0.96, specificity 0.97–0.98, and NPV 0.97-0.98. The best overall result was obtained for classifier U. The metrics were also calculated comparing the experts using expert 1 as ground truth. In comparison to the classifiers most values were somewhat lower for the experts.

**Tab. 1.** Results for the classifiers. Values are averages over folds (±standard deviation). E12: using expert 1 as ground truth and comparing with expert 2; E1: expert 1; E2: expert 2; U: union of expert 1 and 2; I: intersection of expert 1 and 2; Data: indicate which annotation that was used for training and evaluation; ACC: accuracy; B-ACC: balanced accuracy; R-AUC: area under the curve based on receiver operating characteristic; PR-AUC: precision-recall area under the curve; MCC: Matthew’s correlation coefficient; Sens: sensitivity; Spec: specificity; Prec: precision; NPV: negative predictive value. Blue indicates comparison between expert 1 and 2; green indicates results from testing the classifiers; light green indicates results from training the classifiers.

| Data | ACC | B-ACC | R-AUC | PR-AUC | Fo.5 | MCC | Sens | Spec | Prec | NPV |
| --- | --- | --- | --- | --- | --- | --- | --- | --- | --- | --- |
| E12 | 0.93 | 0.74 | 0.49 | 0.50 | 0.60 | 0.53 | 0.52 | 0.97 | 0.62 | 0.95 |
| E1 | 0.94±0.00 | 0.82±0.03 | 0.65±0.05 | 0.60±0.02 | 0.68±0.04 | 0.65±0.03 | 0.68±0.06 | 0.97±0.01 | 0.68±0.06 | 0.97±0.01 |
| E2 | 0.95±0.01 | 0.79±0.05 | 0.58±0.11 | 0.54±0.08 | 0.59±0.14 | 0.57±0.11 | 0.61±0.12 | 0.97±0.01 | 0.59±0.16 | 0.97±0.01 |
| U | 0.94±0.00 | 0.86±0.02 | 0.72±0.05 | 0.65±0.03 | 0.75±0.07 | 0.72±0.04 | 0.75±0.05 | 0.97±0.01 | 0.75±0.09 | 0.97±0.01 |
| I | 0.96±0.01 | 0.76±0.06 | 0.52±0.12 | 0.51±0.07 | 0.53±0.13 | 0.51±0.08 | 0.54±0.13 | 0.98±0.01 | 0.55±0.18 | 0.98±0.01 |
| E1 | 0.96±0.00 | 0.85±0.01 | 0.70±0.03 | 0.67±0.01 | 0.76±0.01 | 0.72±0.01 | 0.73±0.03 | 0.98±0.00 | 0.77±0.03 | 0.97±0.00 |
| E2 | 0.95±0.00 | 0.83±0.04 | 0.66±0.08 | 0.62±0.02 | 0.68±0.04 | 0.65±0.02 | 0.68±0.09 | 0.98±0.01 | 0.68±0.06 | 0.97±0.01 |
| U | 0.96±0.00 | 0.88±0.02 | 0.77±0.04 | 0.70±0.01 | 0.81±0.02 | 0.78±0.01 | 0.79±0.04 | 0.98±0.01 | 0.82±0.04 | 0.97±0.01 |
| I | 0.97±0.00 | 0.83±0.05 | 0.66±0.09 | 0.66±0.03 | 0.71±0.03 | 0.68±0.03 | 0.67±0.10 | 0.99±0.00 | 0.73±0.06 | 0.99±0.00 |

Classifier 1 identified 1,703 EDs, classifier 2 identified 1,431 EDs, classifier U identified 2,228 EDs, and classifier I identified 862 EDs. Out of these false positives, almost all were within the PD population (Tab. 2).

**Tab. 2.** Summary of detected EDs. E12: Comparing the annotations for expert 1 and 2 using expert 1’s as ground truth; C1: classifier based on expert 1’s annotation; C2 classifier based on expert 2’s annotation; CU: classifier based on the union of expert 1 and 2 annotations; CI: classifier based on the intersection of expert 1 and 2’s annotations; TP: true positives; TN: true negatives; FP: false positives; FN: false negatives; ⊂Periodic: Number of EDs detected by the classifiers that were from the population of PDs; ⊄Periodic: Number of EDs detected by the classifiers that were outside the population of PDs.

| Classifier | #EDs–experts | #EDs–classifiers | TP | TN | FP | FN | $\subset$Periodic | $\not\subset$Periodic |
| --- | --- | --- | --- | --- | --- | --- | --- | --- |
| E12 | 1,709 | 1,430 | 886 | 17,155 | 544 | 823 | - | 0 |
| C1 | 1,709 | 1,703 | 1,158 | 17,151 | 545 | 551 | 1,701 | 2 |
| C2 | 1,430 | 1,431 | 898 | 17,450 | 525 | 532 | 1,431 | 0 |
| CU | 2,253 | 2,228 | 1,682 | 16,610 | 542 | 571 | 2,225 | 3 |
| CI | 886 | 862 | 475 | 18,132 | 387 | 411 | 862 | 0 |

The Euclidean distance (‖*x* − *y*‖) between periodic non-EDs (*x*) and EDs (*y*) for the annotations of expert 1 and 2 and the corresponding classifiers were calculated for each channel. The distribution of distances across the electrodes were mostly similar with largest difference for Fp1, Fp2, F8, Cz, Pz, and O1. There were however some differences. Most emphasis was on Fp1, Fp2, Pz, and O1 for expert 1 and on Cz, Pz, and O1 for expert 2. This was also reflected by the classifiers. The classifiers thus preserved the distribution of the distances between periodic non-EDs and EDs compared to the experts (Fig. 4), but the distances increased and more notably so for expert 2 (Tab. 3).

**Tab. 3.** Percentual increase in distance between periodic non-ED and ED when comparing the classifier to expert annotations. The median distances were larger for the classifier annotations (*p* ≪ 0.001; Wilcoxon signed rank test). Top row: electrode. Middle row: percentual increase in distance for expert 1 vs. classifier 1. Bottom row: percentual increase in distance for expert 2 vs. classifier 2.

| Fp1 | F7 | T3 | T5 | Fp2 | F8 | T4 | T6 | F3 | C3 | P3 | O1 | F4 | C4 | P4 | O2 | Fz | Cz | Pz | A1 | A1 |
| --- | --- | --- | --- | --- | --- | --- | --- | --- | --- | --- | --- | --- | --- | --- | --- | --- | --- | --- | --- | --- |
| 9 | 7 | 7 | 6 | 8 | 1 | -3 | 8 | 11 | 10 | 0 | 6 | 5 | 5 | 9 | 16 | 13 | 4 | 1 | 3 | -2 |
| 8 | 10 | 13 | 23 | 9 | 13 | 16 | 20 | 8 | 23 | 15 | 17 | 17 | 18 | 12 | 14 | 13 | 16 | 15 | 18 | 19 |

**Tab. 4.** Interrater agreement according to Cohen’s kappa and Gwet’s AC1 (confidence intervals in parenthesis). The highest value of the respective measure is in bold and underlined. The highest values when at least one expert is included in a comparison are in bold. E1: expert 1; E2: expert 2; C1: classifier based on expert 1; C2: classifier based on expert 2; CU: classifier based on the union of expert 1 and 2; CI: classifier based on the intersection of expert 1 and 2. Comparison between expert is in blue, comparisons between an expert and a classifier are in green, and comparison between classifiers are in yellow.

| Comparison | Cohen’s kappa | Gwet’s AC1 | % agreement |
| --- | --- | --- | --- |
| E1 vs. E2 | 0.53 (0.504–0.549) | 0.92 (0.913–0.922) | 93 |
| E1 vs. C1 | **0.65** (0.630–0.669) | 0.93 (0.927–0.935) | 94 |
| E2 vs. C2 | 0.60 (0.576–0.620) | **0.94** (0.933–0.8940) | 93 |
| E1 vs. C2 | 0.57 (0.548–0.591) | 0.92 (0.920–0.929) | 94 |
| E2 vs. C1 | 0.57 (0.544–0.587) | 0.92 (0.918–0.926) | 95 |
| E1 vs. CU | **0.65** (0.628–0.665) | 0.93 (0.924–0.932) | 96 |
| E2 vs. CU | 0.60 (0.578–0.618) | 0.93 (0.921–0.929) | 94 |
| E1 vs. CI | 0.54 (0.520–0.566) | 0.93 (0.926–0.934) | 94 |
| E2 vs. CI | 0.52 (0.497–0.546) | 0.93 (0.930–0.938) | 94 |
| C1 vs. C2 | 0.67 (0.653–0.692) | 0.94 (0.937–0.945) | 95 |
| C1 vs. CU | **0.82** (0.802–0.831) | **0.96** (0.959–0.965) | 97 |
| C2 vs. CU | 0.72 (0.701–0.737) | 0.95 (0.944–0.951) | 96 |
| C1 vs. CI | 0.67 (0.646–0.687) | 0.95 (0.943–0.951) | 95 |
| C2 vs. CI | 0.70 (0.680–0.722) | **0.96** (0.956–0.962) | 96 |
| CU vs. CI | 0.67 (0.654–0.694) | 0.95 (0.942–0.949) | 95 |

**Fig. 3.**
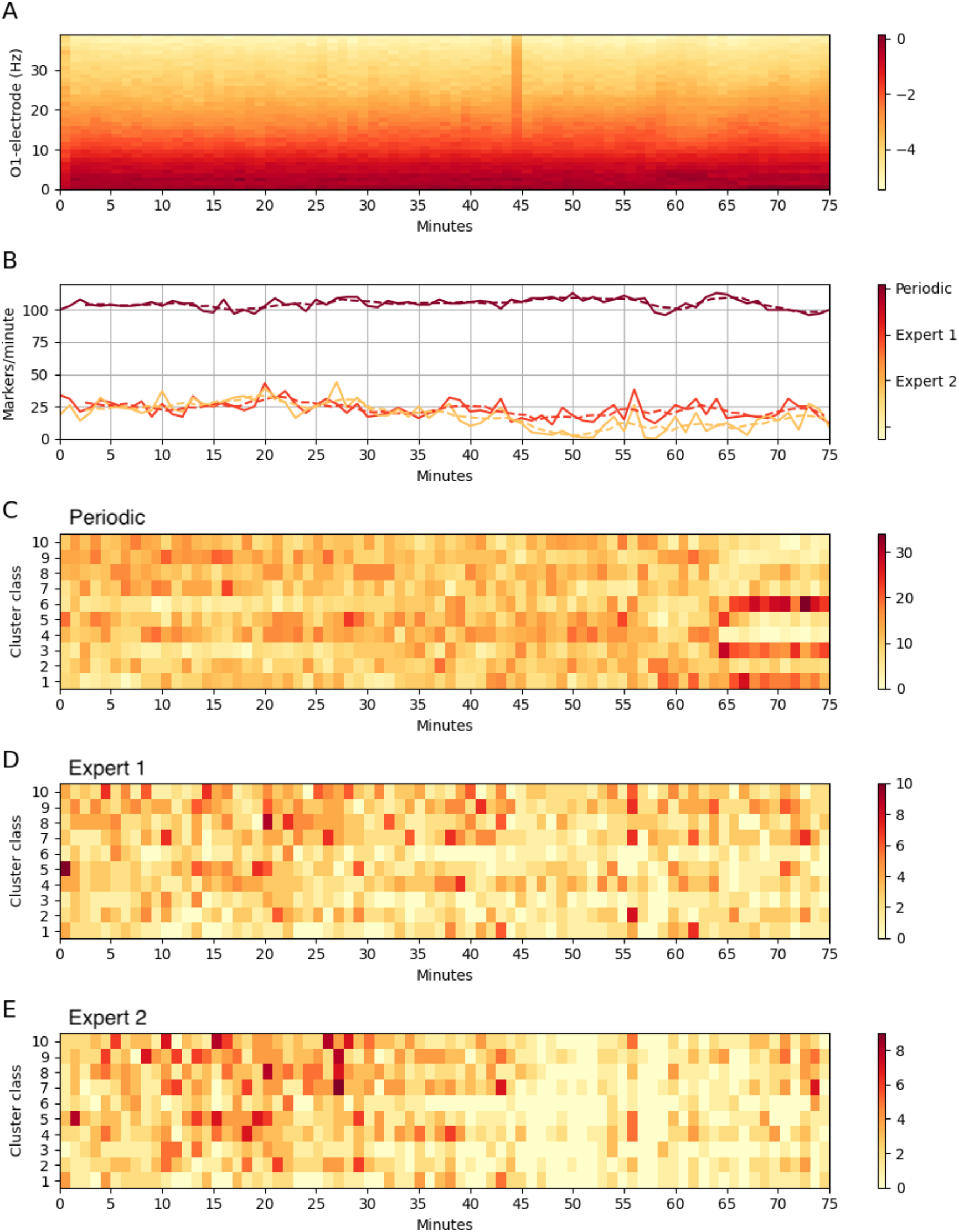
Overview of the data and results as a function of time. (A) Frequency spectrum below 40 Hz for the O1-electrode. The color bar indicates the mapping of colors to the logarithm of the mean absolute amplitude per minute. (B) Number of markers per minute for the annotations. Dashed lines are five-minute moving average. (C) Distribution of periodic markers with respect to cluster classes. The color bar indicates the mapping of colors to the number of markers per minute. (D) Distribution of expert 1’s markers with respect to cluster classes. The color bar indicates the mapping of colors to the number of markers per minute. (E) Distribution of expert 2’s markers with respect to cluster classes. The color bar indicates the mapping of colors to the number of markers per minute.

**Fig. 4.**
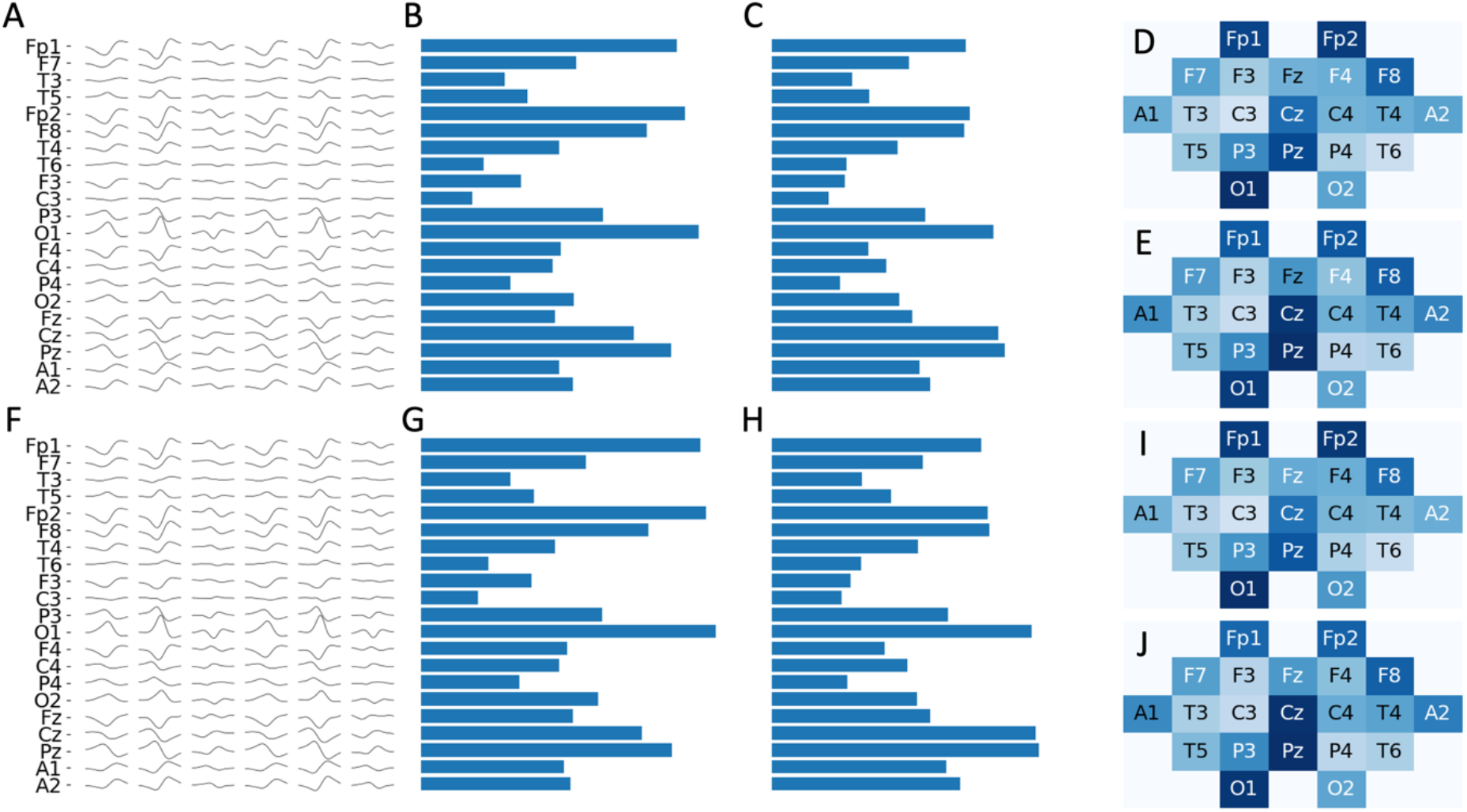
Euclidean distances between the averages for periodic non-EDs and EDs. In the field plots D, E, I, and J the color scale is from white (0) to dark blue (max distance). (A) First potential: average ED by expert 1; second potential: average non-ED by expert 1; third potential: average difference by expert 1; fourth potential: average ED by expert 2; fifth potential: average non-ED by expert 2; sixth potential: average difference by expert 2. (B) Euclidean distances per electrode for the averages of expert 1. (C) Euclidean distances per electrode for averages of expert 2. (D) Field plot for the Euclidean distances for averages of expert 1. (E) Field plot for the Euclidean distances for the averages of expert 2. (F) First potential: average ED by classifier 1; second potential: average non-ED by classifier 1; third potential: average difference by classifier 1; fourth potential: average ED by classifier 2; fifth potential: average non-ED by classifier 2; sixth potential: average difference by classifier 2. (G) Euclidean distances per electrode for the averages of classifier 1. (H) Euclidean distances per electrode for averages of classifier 2. (I) Field plot for the Euclidean distances for averages of classifier 1. (J) Field plot for the Euclidean distances for the averages of classifier 2.

### 4.3. Interrater agreement

The Cohen’s kappa value for the experts was 0.53 and the kappa values when comparing the experts to the classifiers were 0.52–0.65 (Tab. 4). There was thus a moderate and comparable agreement between the experts and between experts and classifiers. The kappa values when comparing the classifiers with each other were 0.67-0.82 (Tab. 4), mostly corresponding to a substantial agreement.

Using Gwet’s AC1 produced a different result with almost perfect agreement. For the experts, the value was 0.92 and when comparing the experts to the classifiers the values were 0.92–0.94 (Tab. 4). When comparing the classifiers (trained with different annotations) to each other, the values were 0.94–0.96 (Tab. 4).

The percent agreement comparing the experts was 93 percent, experts compared to the classifiers 93–96 percent, and comparing the classifiers 95–97 percent (Tab. 4).

### 4.4. Cluster analysis

A cluster analysis was performed for the annotations of expert 1 and 2 and the corresponding classifiers. The cluster encoder produced distinct clusters of the PDs and the automatic cluster identification works (Fig. 5). Unfortunately, there were three examples that produces zero output (Fig. 5; cluster 11).

**Fig. 5.**
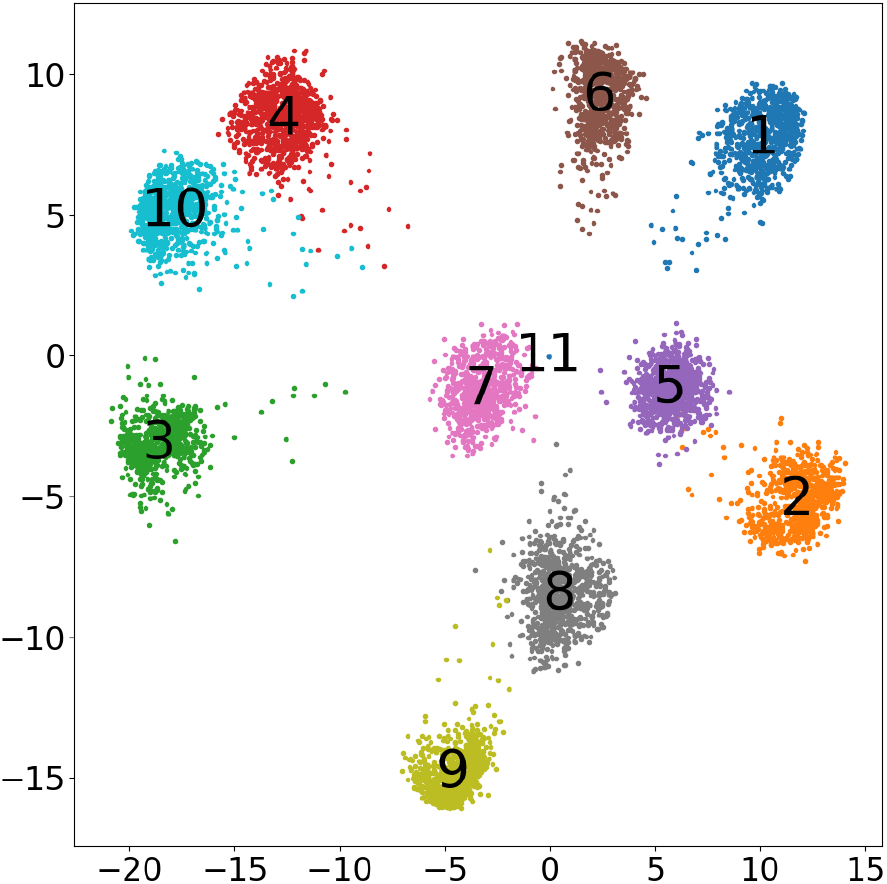
Automatically color-coded clusters obtained from the cluster encoder. Each dot represents one EEG segment of size 21×64 (channels x time steps).

Most of the discharges identified as EDs aggregated, and the distribution of aggregates were similar for experts and classifiers (Fig. 6 and Fig. 7). In most instances, differences concerned data points that were close. There were also indications that the experts assessed more consistently different relative each other for some discharges, and that this affected the learning of the classifiers (Fig. 8). For example, in cluster 2 and 9, expert 1 and 2 tended to select from opposing sides of the clusters. In cluster 2, expert 1 selected more compared to expert 2, while the opposite was true for cluster 9. There were also differences between the experts and the corresponding classifiers, e.g., in cluster 1, the classifier was more prone to identify EDs compared to expert 1 while the opposite was true for expert 2 and the corresponding classifier. The experts only identified a few scattered EDs in cluster 10, only one of these were common to the experts. Expert 2 also identified a few in cluster 4, where expert 1 did not identify any EDs. The classifiers did not identify any EDs in any of the two clusters.

**Fig. 6.**
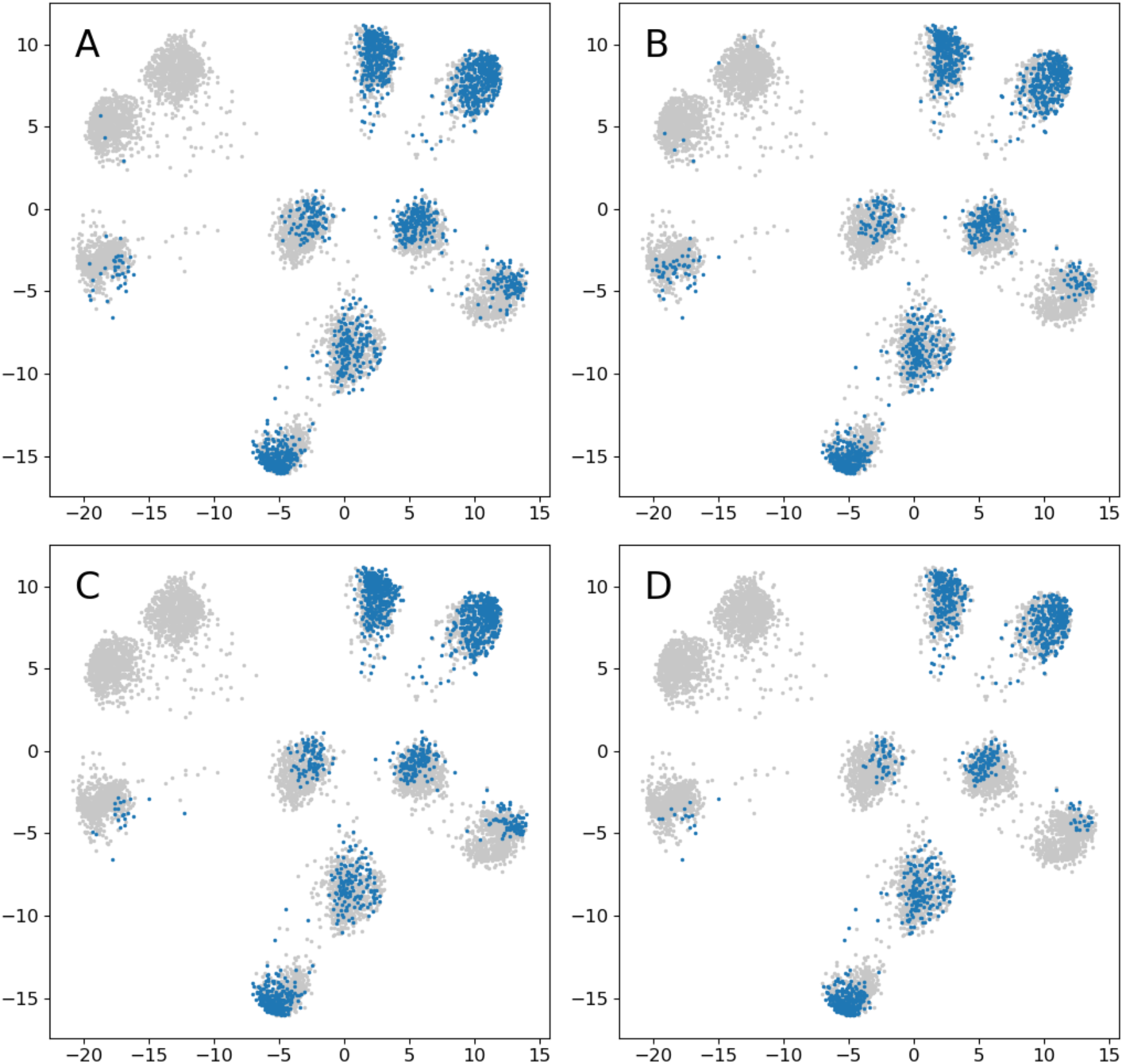
Distribution of all the annotations. Blue: ED; grey: non-ED. (A) Expert 1. (B) Expert 2. (C) Classifier 1. (D) Classifier 2.

**Fig. 7.**
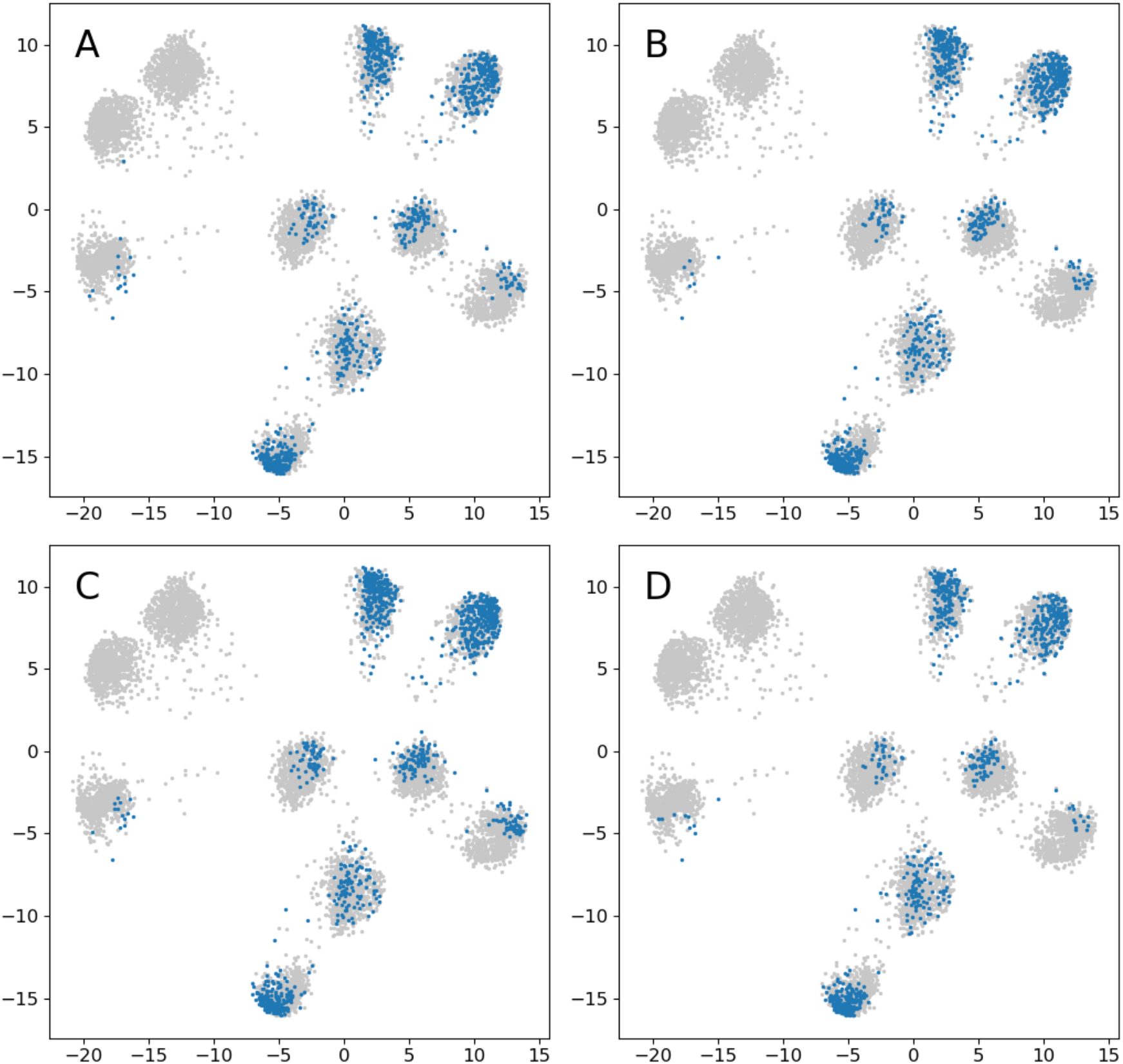
Comparison of intersections between annotations. Blue: ED; grey: non-ED. (A) Expert 1 and expert 2. (B) Classifier 1 and classifier 2. (C) Expert 1 and classifier 1. (D) Expert 2 and classifier 2.

**Fig. 8.**
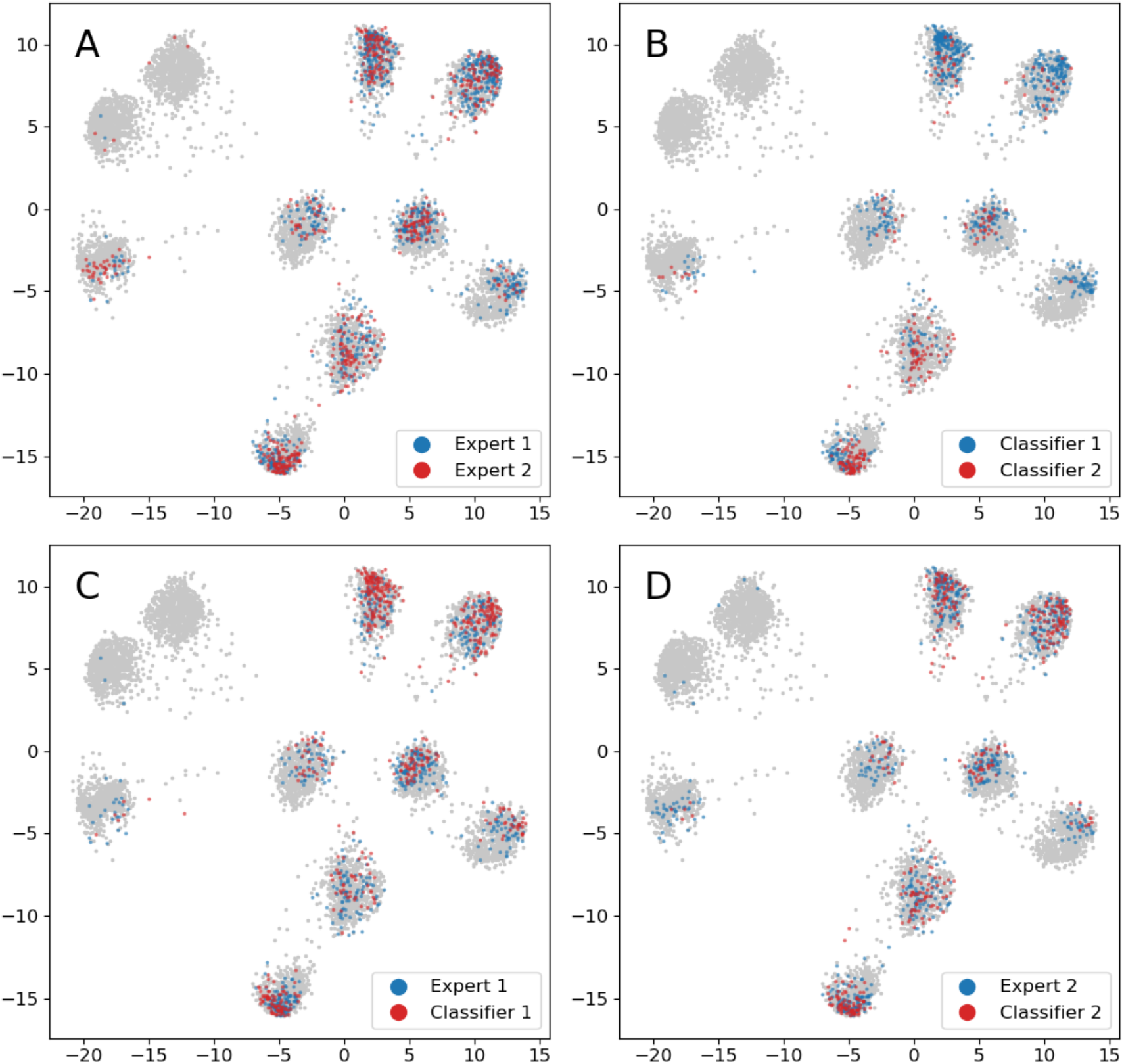
Comparison of annotations unique to one of the raters. (A) Expert 1 (blue) and expert 2 (red). (B) Classifier 1 (blue) and classifier 2. (red) (C) Expert 1 (blue) and classifier 1 (red). (D) Expert 2 (blue) and classifier 2 (red).

Averages of the discharges for each cluster are presented in Appendix 9.2. It is difficult to draw any major conclusions from the averages. There was some variation in the electric fields of the clusters. For the O1 electrode, averages of EDs were of higher amplitude and of sharper morphology compared to non-EDs. The overall averages of cluster 4 and 10 had a blunter morphology compared to the other clusters, in line with that non to few were identified as EDs in the annotations.

## 5. Discussion

### 5.1. Agreement scores

For the percent agreement, Cohen’s kappa and Gwet’s AC1, comparing classifiers achieved the highest agreement, followed by experts compared to classifiers, while comparing the experts produced the lowest scores. As expected, agreement scores were higher when using Gwet’s AC1, where differences in agreement were small.

The two experts identified 1,709 and 1,430 EDs, respectively, but only agreed on 886. According to the often-used interpretation by Landis and Koch (1977), Gwet’s AC1 indicate an almost perfect agreement for the experts. This is not a reasonable interpretation and support the conclusion by Vach and Gerke (2023) that this ordinal scale should not be used for Gwet’s AC1.

### 5.2. Disagreement – similar or different?

Although there were differences between the annotations, the cluster analysis showed that annotated examples tended to aggregate, i.e., were similar, and that the different annotations showed similar distributions of aggregates. The cluster analysis also showed that differences in expert annotation to some extent induced corresponding differences for the classifiers (e.g., Fig. 8 A and B; clusters 2, 3, and 9). However, as the clustering method is new, it is uncertain how strong conclusions that can be made, and as discussed below, it must be further and more systematically evaluated.

Jing et al. (2019) suggest that experts rate EDs based on the same internal model and that differences in rating mostly are due to differences in their thresholds for scoring EDs. Assuming that: 1) the experts use the same model for classifying, 2) the only significant difference between them is a threshold, and 3) that the thresholds are simple and stable; then most of the examples classified as EDs by the expert with higher threshold should be a subset of the other expert’s selected examples (Fig. 9 A). This was not the case here since each expert has a subset of significant number of examples that were uniquely scored as EDs by the respective expert (Fig. 9 B).

**Fig. 9.**
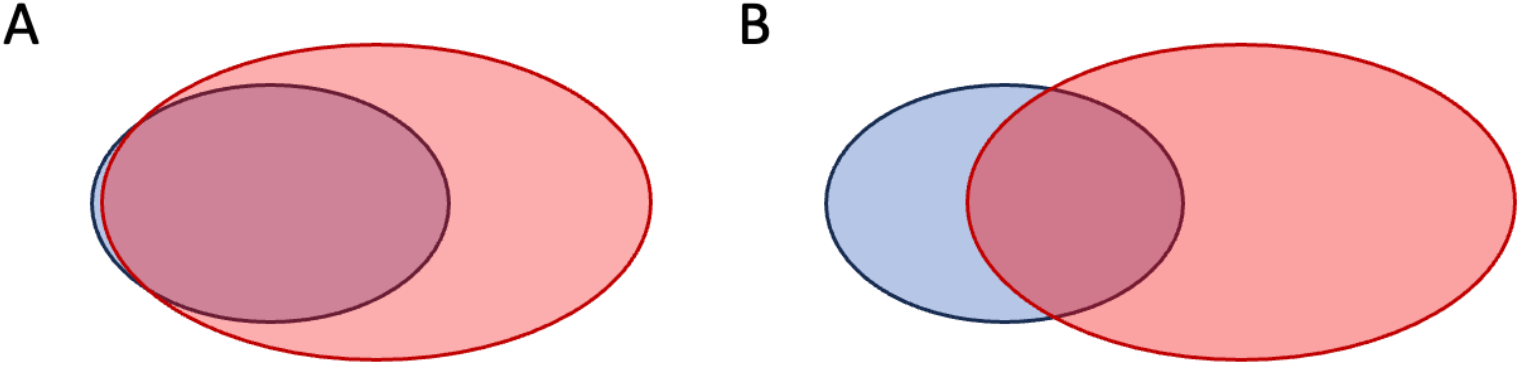
Illustration of two experts’ annotations where one expert (red) has a lower scoring threshold, i.e., score more, compared to the other expert (blue). (A) In this case almost all of the high threshold expert is a subset of the low threshold expert. This is consistent with them using the same model and one (blue) having a higher scoring threshold. (B) In this case there is an overlap between the scoring, but each expert also has their respective scored subset that are non-overlapping. This would imply either that they use different models for scoring or that they have different sensitivities along different dimensions of a multi-dimensional threshold.

The distances between EDs and non-EDs (Fig. 1) show that the annotation of expert 1 seemed to emphasize the dipole O1, Fp1, Fp2, while the annotation of expert 2 emphasized the peak Cz, Pz, and O1. This was reflected in how the classifiers performed. The result thus indicates that the experts to some extent rate differently and that this affects the learning of the classifiers.

Interpreting the cluster analysis cautiously, part of the data seems to be assessed more differently by the experts (e.g., Fig. 8 A; clusters 2, 3, and 9). Assuming that scoring is based on a common model, this could imply a multidimensional threshold, and the more different assessments may represent dimensions where the experts had a more consistent difference in threshold. Part of the data that has been scored differently seem to be around more similar data points, which could imply that the thresholds are closer but vary over time, i.e., intrarater variability. In this case, it can thus vary which expert is most sensitive to scoring, and this could contribute to different but similar annotations.

### 5.3. The classifiers

There are several factors the classifiers cannot learn. For instance, the annotation process consumed several hours, and fatigue may have affected the thresholds. This is a form of external factor, not encoded in the EEG. The experts were instructed to assess each candidate discharge independently, but recently scored material may still affect the threshold, e.g., if there is a period where discharges are blunter, the threshold may be lowered for scoring a discharge as epileptiform. Of course, there may also be long-term biological dependencies in the data that are clinically relevant and influence how the experts detect EDs. The classifiers cannot learn variation caused by external factors, or long-term dependencies since this type of classifier’s assessment is independent of the location in the data. It is therefore further speculated that the classifiers learn some form of average threshold.

In a practical setting, how to balance between false positives and negatives will depend on the application. For the alternatives tested, none produced a perfect classifier, and since the aim of the study was to compare annotations, it was decided to train the classifiers toward producing roughly the same amount of EDs as the annotations they were trained on. There were limitations in creating the classifiers, e.g., they had a very basic architecture, no extensive hyperparameter tuning was performed, and the data size used to train them was relatively small. A problem during this work was the large variation in validation values (accuracy, false positive and negative predictions) from iteration to iteration during training. This made it difficult to predict how the resulting classifiers would perform when tested (it was hard to aim for something). The weights of the classifiers were initialized with the same randomized values, but there were other sources of randomness that occurred during training, e.g., randomization of the training data, that could have contributed to a variation in the final results. In a larger study, this variation could be assessed by training multiple classifiers on the same data. There was some correlation between the performance metrics (Tab. 1) and the number of PDs in each annotation with higher values for larger number of PDs. It may be possible to tune the classifiers to classify more aligned to the experts, but the relatively small dataset will set a limit to what is possible to achieve. Interpreting the (balanced) performance metrics in a traditional way, the classifiers would be assessed as performing relatively poorly. However, when comparing the experts to each other using the same metrics, the classifiers had a better performance. Almost all false positive EDs identified by the classifiers were within the PD population. Part of the false positives would possibly be accepted as EDs, and part of the false negatives would possibly be accepted as rejected EDs by experts in a Turing test. Regardless of room for finetuning the classifiers and using larger data sizes, it is speculated that the classifiers probably cannot learn to reproduce the experts’ annotations exactly, and that intrarater variability at least to some degree could contribute to this.

### 5.4. The EEG data

The aim of the study was not to settle the question of what constitutes EDs, but to study how rating varies between experts and how this affects the learning by classifiers. Only one EEG was used for this study. There were several reasons for this choice. The EEG was longer than a standard exam, so it would provide more training data for the classifiers while still being reasonable for an expert to annotate in one session. The chosen exam represented a focal status epilepticus and so would contain relatively large amounts of candidate EDs. This would also increase the chance of training good classifiers based on a smaller amount of EEG. The PDs had a variation in morphology, but relatively similar electric fields. Hence, there would be a combination of a large but still limited variation that would provide more extensive testing of the scoring thresholds of the experts. The times to perform the annotations were shorter than expected and it seem possible to use recordings of longer duration.

### 5.5. The cluster method

The basis for the cluster encoder has been evolving over an extended period of time. The current version of the encoder has several limitations. Similar to k-means clustering, the number of clusters has to be decided beforehand. Furthermore, there is no guarantee that all clusters will be utilized. The architecture may cause zero output for some data, and this is a more serious problem. This is represented by cluster 11 in the results. The number of clusters can affect the degree of zero output. The loss function will promote a uniform distribution of examples across the clusters. This may cause blending of categories in clusters if the training data have skewed prevalence of categories. In addition to the number of clusters, there are several new parameters that must be chosen (e.g., neighbors, sigmoid distribution, weights for loss function).

For different data tested during the development, the best results have been produced using 5–15 clusters and a low number of neighbors (1–10). In many instances, a problem has rather been that clusters tend to be almost point-like, compared to the more confluent character produced by previous versions of cluster encoders. The slope parameter *S* of the sigmoid distribution may be less important but using a steep slope (larger value) might promote clusters to expand. Similarly, but probably more important, the bias parameter *b* may also cause clusters to expand by using larger values. The sigmoid distribution can produce very small values. It may be the case that differences in smaller values may not convey information about rank, but rather be due to machine precision. Furthermore, in practice, to avoid division by zero, a lower limit must be set for the distribution. If the sigmoid distribution produce values below this limit, the corresponding data points will be modeled as equidistant. Using the binary cross-entropy may therefore not be a good option, but the impression was that it improved the results.

Possibly, the clustering and separation is artificially good, i.e., the architecture forces the data into well separated clusters. Furthermore, there were a few data points color coded as belonging to one cluster but are closer to another (e.g., cluster 2 and 5; Fig. 5). It seems more likely that this color coding was an effect of the loss function promoting a uniform distribution of examples per cluster rather than coding for similarity. However, for most data, the color coding seemed reasonable relative the similarity.

### 5.6. Conducting a larger study

In this pilot study, only two experts annotated the data, and the experts were also members of the research group. Studies by Halford et al. (2017) and Bagheri et al. (2017), imply that around ten experts can yield a stable group consensus, and is a reasonable size for a larger study. Ideally, a larger study should include experts from different centers and non-members of the research group. In addition, intrarater agreement could be assessed by having the experts perform the annotation twice on separate occasions. The EEG data and performed annotations in this pre-study can be used to optimize the classifiers to perform more aligned to the experts. New EEG data can then be used to assess the larger expert group. The new data should be similar, but if possible have longer duration.

## 6. Conclusions

The different measures of interrater agreement disagree. However, the general conclusion, including the cluster analysis, is that experts and classifiers rate similar discharges as EDs. From a practical point of view regarding the applicability of machine learning to EEG assessment in clinical practice, these techniques may well come to contribute to more uniform evaluations, possibly closer to the biological basis of the assessed signals. The small sample size of the study makes the results uncertain and strong claims cannot be made.

## Data Availability

EEG data were taken from https://isip.piconepress.com/projects/tuh_eeg/

## 7. Acknowledgements

We are very grateful to dr Saad Nagi for supporting this work. Grants were received from Region Östergötland (RÖ-974228, RÖ-962769, RÖ-941377, RÖ-986017, LIO-936176, and RÖ-941359). A.E. was supported by the ITEA3/VINNOVA funded project (AS-SIST).

## 8. Author Contributions

M.S.: conceptualization, formal analysis, investigation, methodology, project administration, resources, software, visualization, writing—original draft, writing—review and editing. A.E.: supervision, writing—review and editing. M.T.: supervision, writing—review and editing.

## 9. Appendix

### 9.1. Examples of discharges

All discharges were first sorted according to increasing amplitude in the O1-electrode and twenty-one evenly spaced discharges from this order were then plotted (Fig. 10; Fig. 11; Fig. 12).

#### 9.1.1. Periodic discharges

**Fig. 10.**
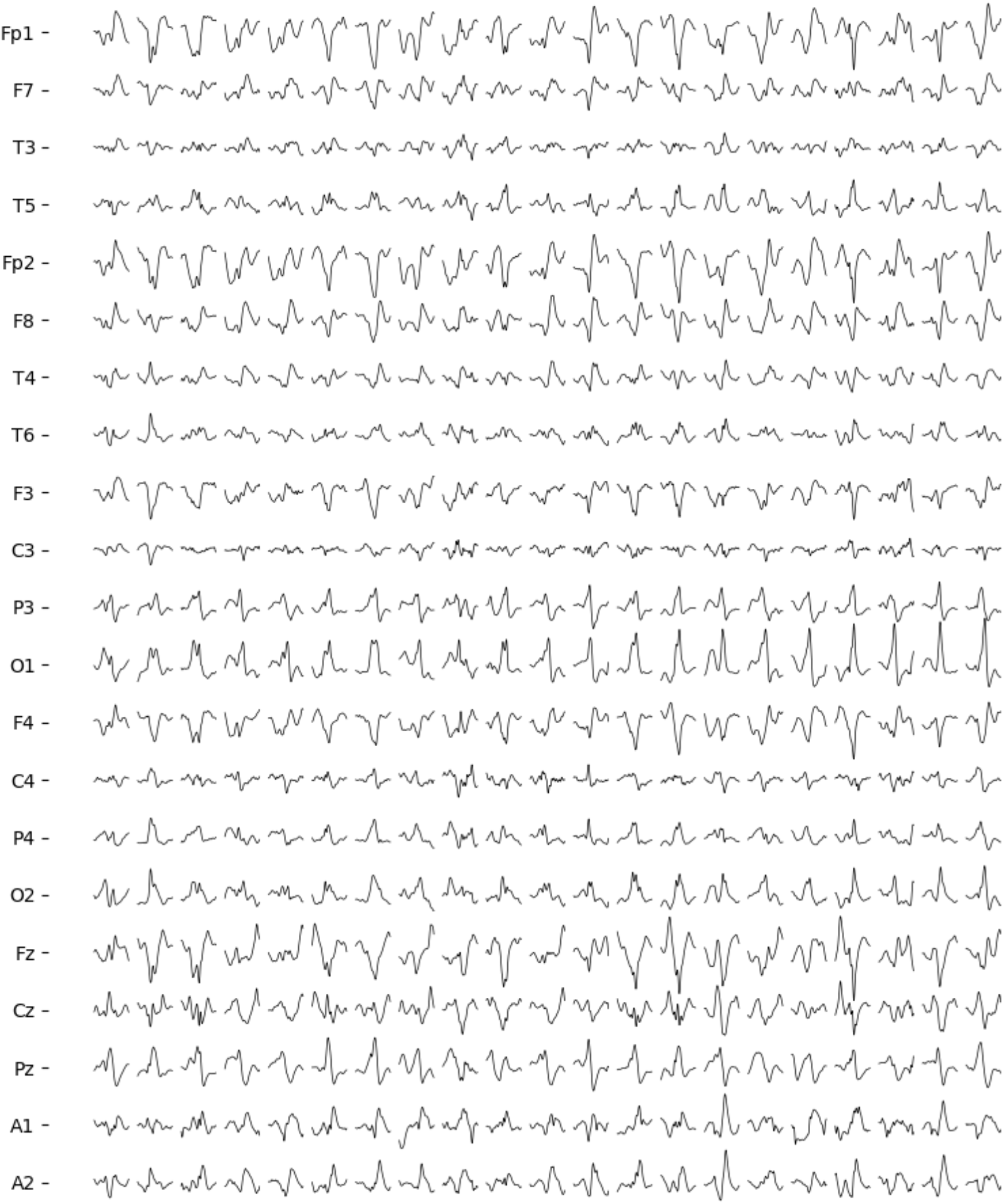
Twenty-one examples of waveforms annotated as periodic discharges. Electrodes referenced to the common average.

#### 9.1.2. EDs by Expert 1

**Fig. 11.**
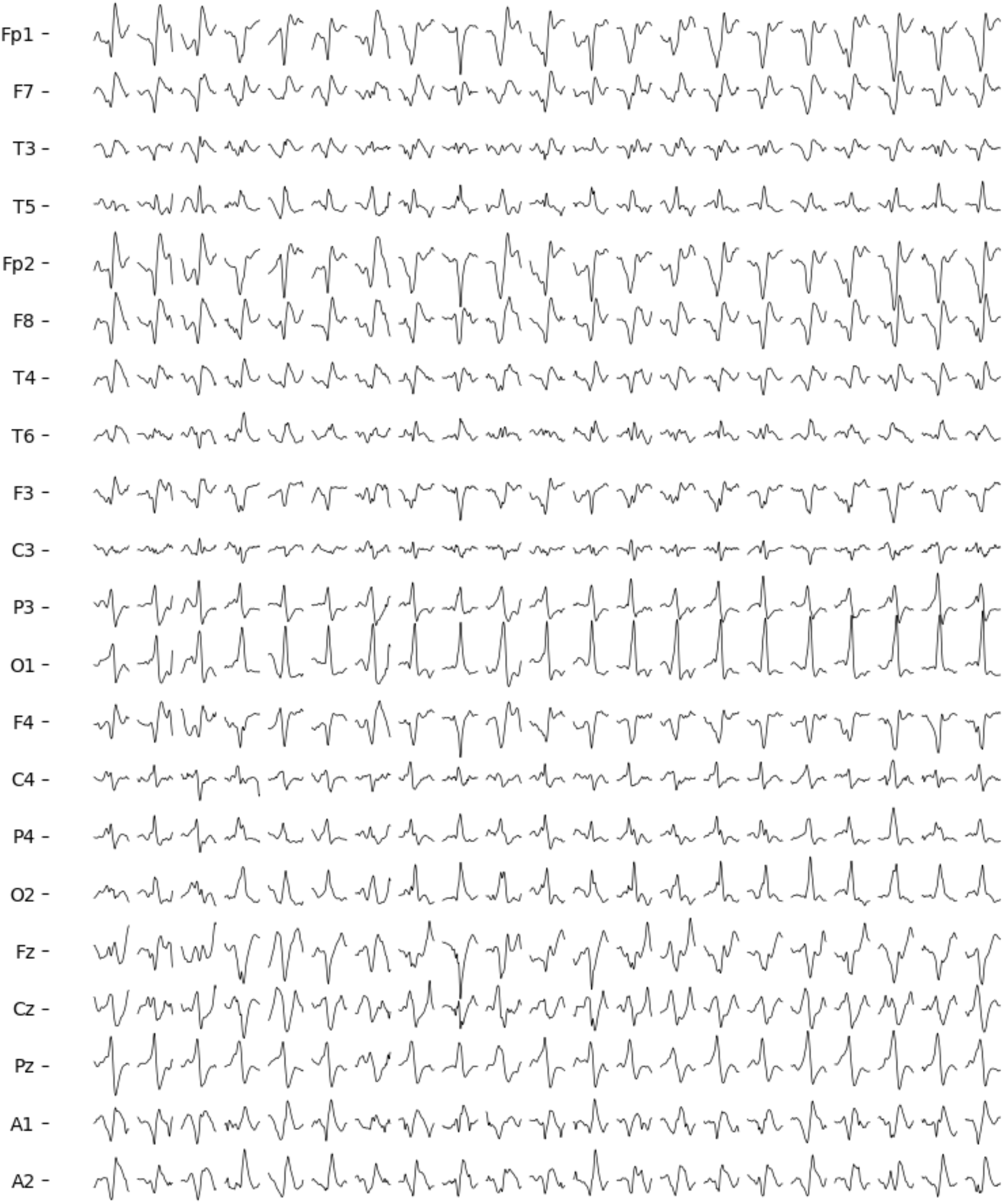
Twenty-one examples of waveforms annotated as epileptiform discharges by expert 1. Electrodes referenced to the common average.

#### 9.1.3. EDs by Expert 2

**Fig. 12.**
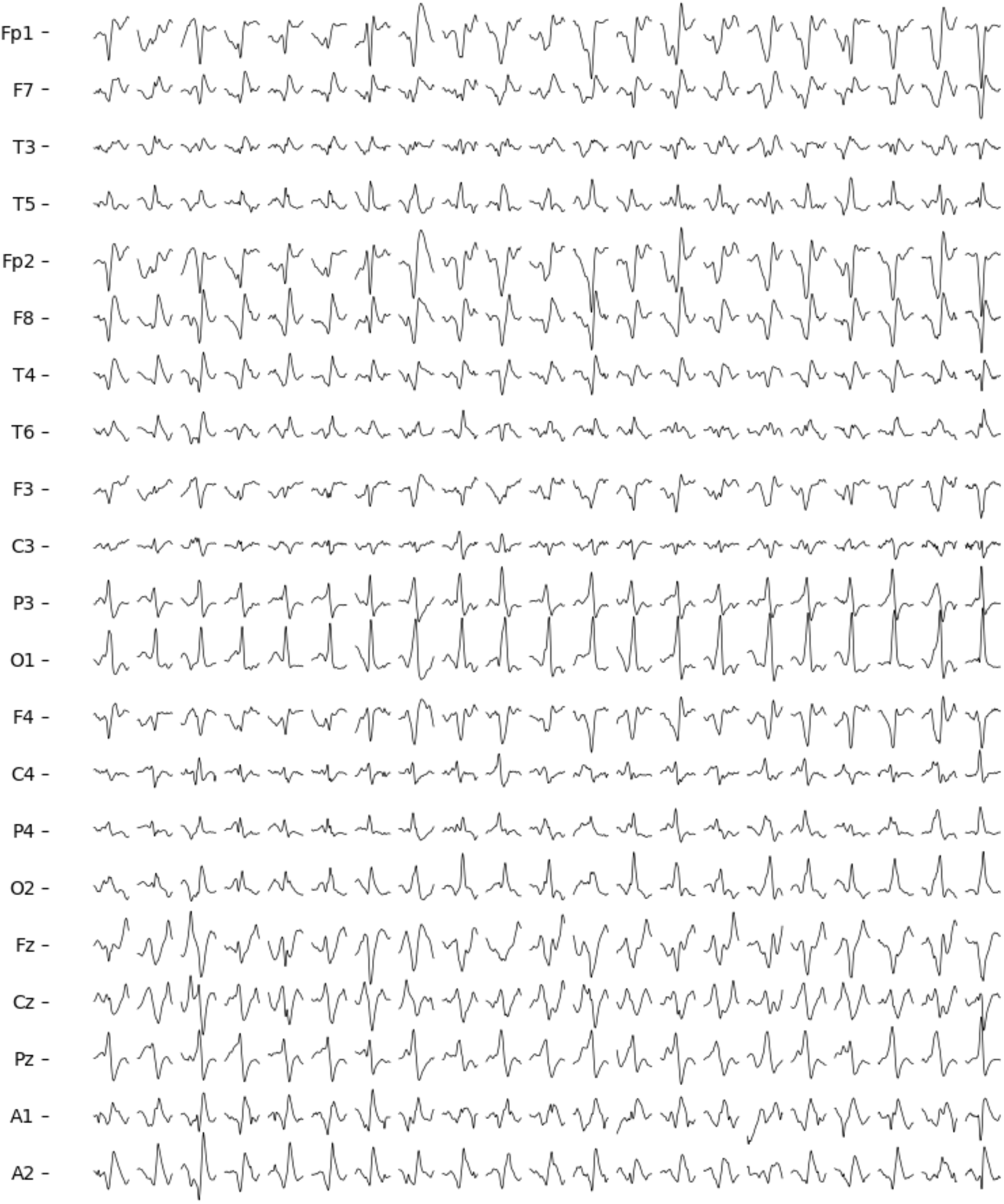
Twenty-one examples of waveforms annotated as epileptiform discharges by expert 2. Electrodes referenced to the common average.

### 9.2. Average discharges of clusters

Averages for the clusters were calculated for both experts and the corresponding classifier. In the figures below, the same color code as in Fig. 5 was used. For each cluster the following averages were calculated of: 1) all discharges, 2) non-EDs of the expert, 3) non-EDs of the classifier, 4) EDs of the expert, and 5) EDs of the classifier.

### 9.3. Cluster encoder

Cluster analysis was performed using a further development of parametric t-distributed stochastic neighbor embedding (t-SNE) that allowed for automatic identification of different clusters.

In t-SNE, a low-dimensional representation (LDR) can be generated of a high-dimensional representation (HDR) of data (van det Maaten and Hinton, 2008). This is accomplished by matching probabilities of data points being neighbors in both the HDR and LDR. The HDR is assumed to have a normal distribution, the LDR a t-distribution, and the distributions are match using the Kullback-Leibler divergence. In parametric t-SNE, this is implemented as a fully connected neural network (van der Maaten, 2009). In recent work, this was adapted to EEG to create cluster encoders (Svantesson et al., 2023). The encoders consisted of convolutional layers that produced a new HDR. Instead of following a normal distribution, the corresponding neighbor distribution was assumed to follow a two-value function based on ranked distances. That is, data points below a certain rank threshold were regarded as neighbors and given a high probability, while those above the threshold were given a low probability.

In this work the method was further developed. The encoder (Fig. 15; Tab. 5) produced two HDRs, *Z* and *C*, that were both matched to a LDR *Y*, as well as to each other. The two-value function used previously only model whether points are neighbors or not. To additionally provide information of the rank, i.e., a proxy value of distance, a sigmoid probability distribution was used instead. The Kullback-Leibler divergence emphasize high probabilities, i.e., close data points. To possibly model distant data points better, the binary cross-entropy was used.

**Tab. 5.** Characteristics of the parts of the encoder. Number of layers refer to convolutional or connected layers. Colors indicate parts forming units in the practical implementation and their corresponding number of parameters. All parameters were trainable.

| Part | Layer type | Number of layers | Size (number of filters or nodes) | Activation | Normalization | Regularization | Number of parameters |
| --- | --- | --- | --- | --- | --- | --- | --- |
| 1 | Convolutional and Max pooling | 6 | Layer 1-5: 64, 128, ..., 1024; layer 6: 128 | SELU | - | L2 | 2,353,408 |
| 2 | Channel-wise connected | 6 | Alternating 512–128 | ReLU | Layer normalization | L2 | 26,148,426 |
|  | Locally connected | 6 | Alternating 512–128 | ReLU | Layer normalization | L2 |  |
| 3 | Fully connected | 6 | Alternating 2688–512 | ReLU | Layer normalization | L2 |  |
| 4 | Fully connected | 6 | Alternating 2688–512 | ReLU | Layer normalization | L2 |  |
| 5 | Locally connected | 6 | Alternating 512–32 | ReLU | Layer normalization | L2 | 1,999,746 |
| Total number of parameters |  |  |  |  |  |  | 30,501,580 |

**Fig. 13.**
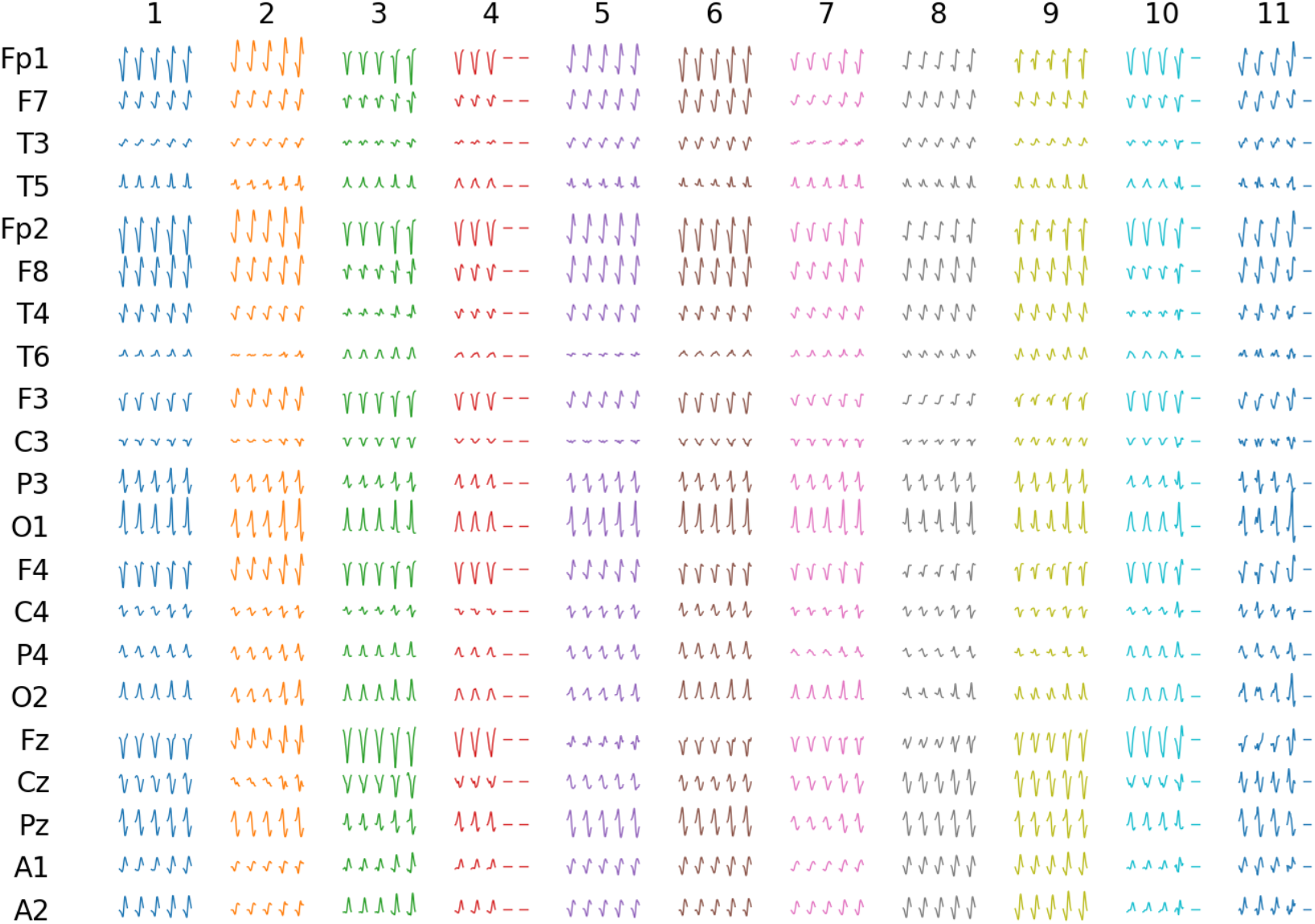
Average discharges for expert 1 and classifier 1. Clusters are indicated by number at the top and the color code is according to Fig. 5. For each cluster there are five averages plotted: 1) the total average of the cluster, 2) average non-ED of for expert, 3) average non-ED for classifier, 4) the average ED for the expert, and 5) the average ED for the classifier.

**Fig. 14:**
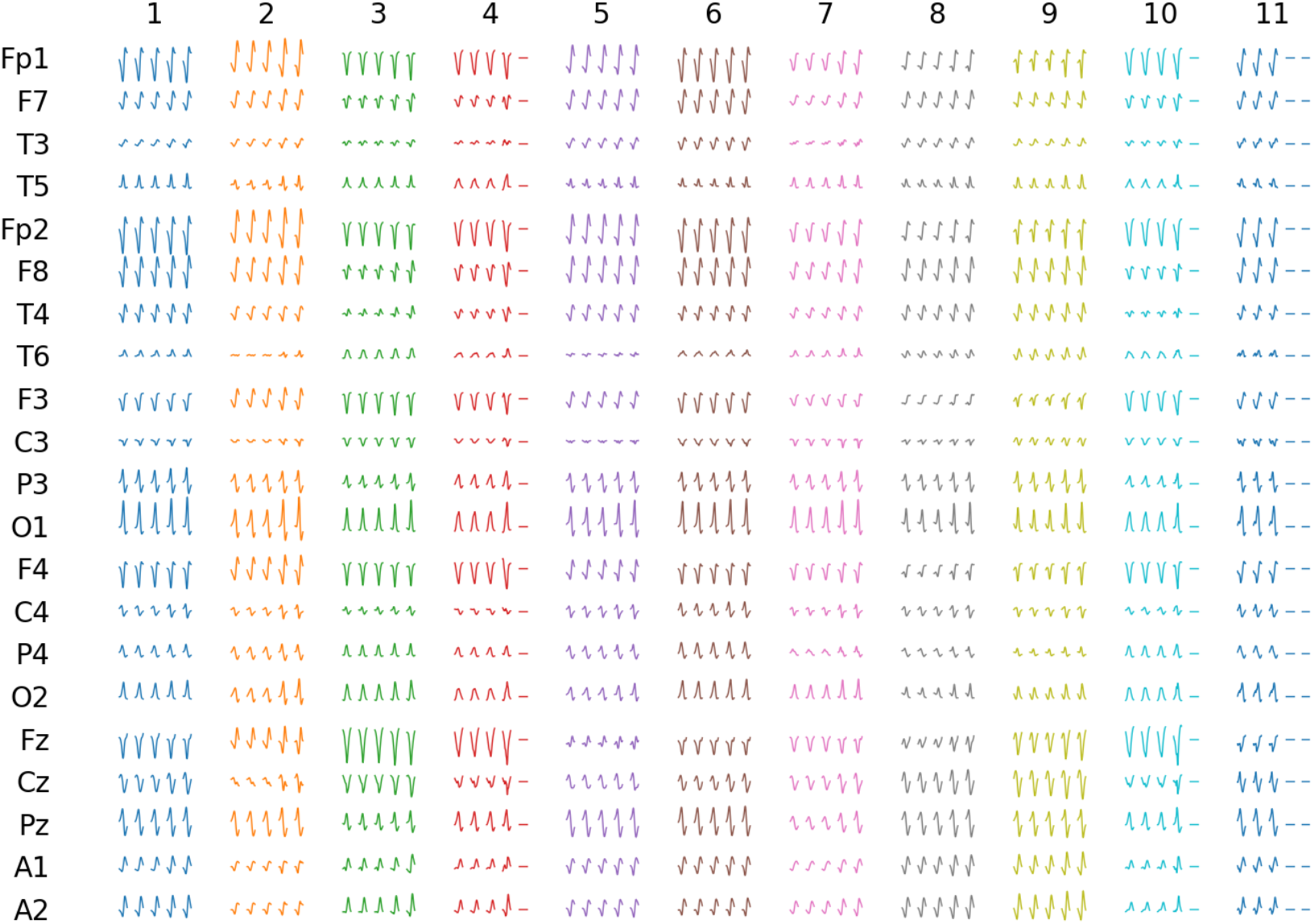
Average discharges for expert 2 and classifier 2. Clusters are indicated by number at the top and the color code is according to Fig. 5. For each cluster there are five averages plotted: 1) the total average of the cluster, 2) average non-ED of for expert, 3) average non-ED for classifier, 4) the average ED for the expert, and 5) the average ED for the classifier.

**Fig. 15.**
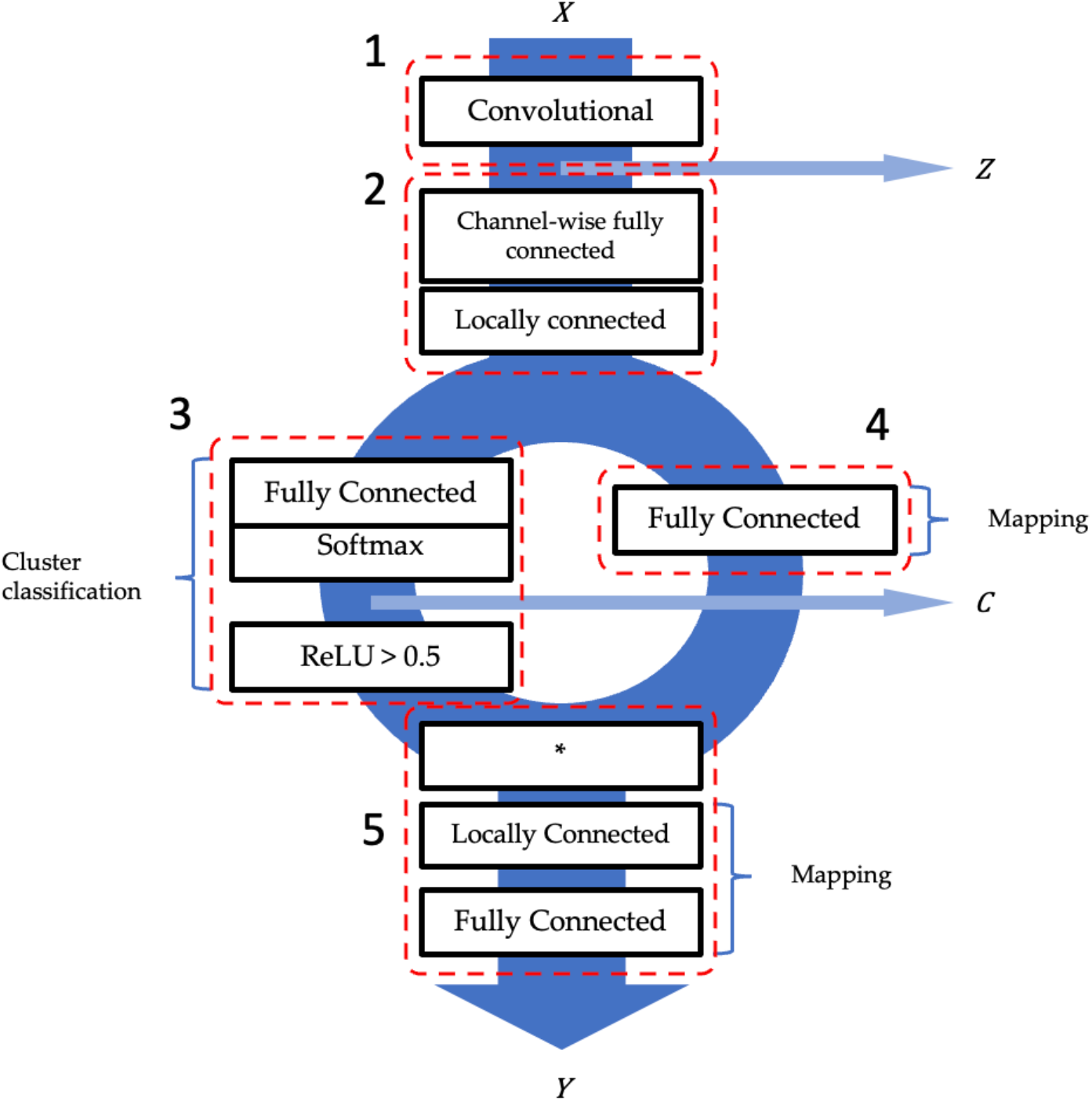
Cluster encoder architecture. Its five parts indicated with numbers 1–5. (1) A series of convolutional layers produced a high-dimensional representation (HDR) ***Z*** of EEG. (2) A series of connected layers. In the first section, the same fully connected layers were applied independently to each channel. In the second section, each layer has nodes that were separately connected per channel. (3) Fully connected layers, analyzing all channels together to determine cluster assignment. It ended in a fully connected layer followed by a softmax activation and then a ReLU activation with a threshold of 0.5. Another HDR ***C*** was extracted before the ReLU activation. (4) Fully connected layer that analyzed all channels together to assess mapping into lower dimensions. (5) Results from part 3 and 4 were first multiplied together. The result from part 3 would then select part of the result from part 4 to preserve information of cluster assignment. This was then further analyzed by locally connected layers, i.e., data corresponding to the respective cluster were analyzed separately. The part ended in a fully connected layer of two nodes, producing the low-dimensional representation ***Y***.

The cluster encoder consisted of five main parts (Fig. 15; the corresponding parts indicated by numbers in the figure):

1. The first part of the cluster encoder was a section of convolutional and max pooling layers, similar to the classifiers, which produced the representation *Z*. Convolutions were performed channel-wise and the size of *Z* was 21×64.
2. The second part started with a section of fully connected layers analyzing channel-wise, which was followed by a section of locally connected layers. The channels were kept separate. The first section thus performed a more general waveform analysis, while the second section performed analysis adapted to each channel. The basic size was kept 21×64.
3. The third part received input from the second part and consists of fully connected layers which ended in a layer with the number of nodes equal to the number of clusters, and this was followed by a softmax which produced the representation C. This part was then ended by a ReLU with a threshold of 0.5. The idea is that the combination of a softmax function followed by a ReLU with threshold 0.5 can at most generate a value (0.5–1) through one node. The output size from this part was 10 and it was used to produce automatic cluster identifier (numbers or color code).
4. The fourth part also received input from the second part and consisted of fully connected layers. The output size from this part was 10×32.
5. The fifth part received input from the third and fourth part. These inputs were first multiplied and then processed by locally connected layers and ended in a fully connected layer with two nodes which produced the LRD Y. In the locally connected layers, the basic size was the number of clusters times the size of the locally connected layers, which for this work was 10×32.

Locally and fully connected sections of parts two to five had the general structure of blocks composed of a series of layers (Fig. 16): connected layer, ReLU, connected layer, ReLU, add residual, and layer normalization. The first connected layer either expanded or contracted the input size to 512, while the second connected layer restored it to the original size. The block is reminiscent of the feedforward network of the transformer (Vaswani et al., 2017).

**Fig. 16.**
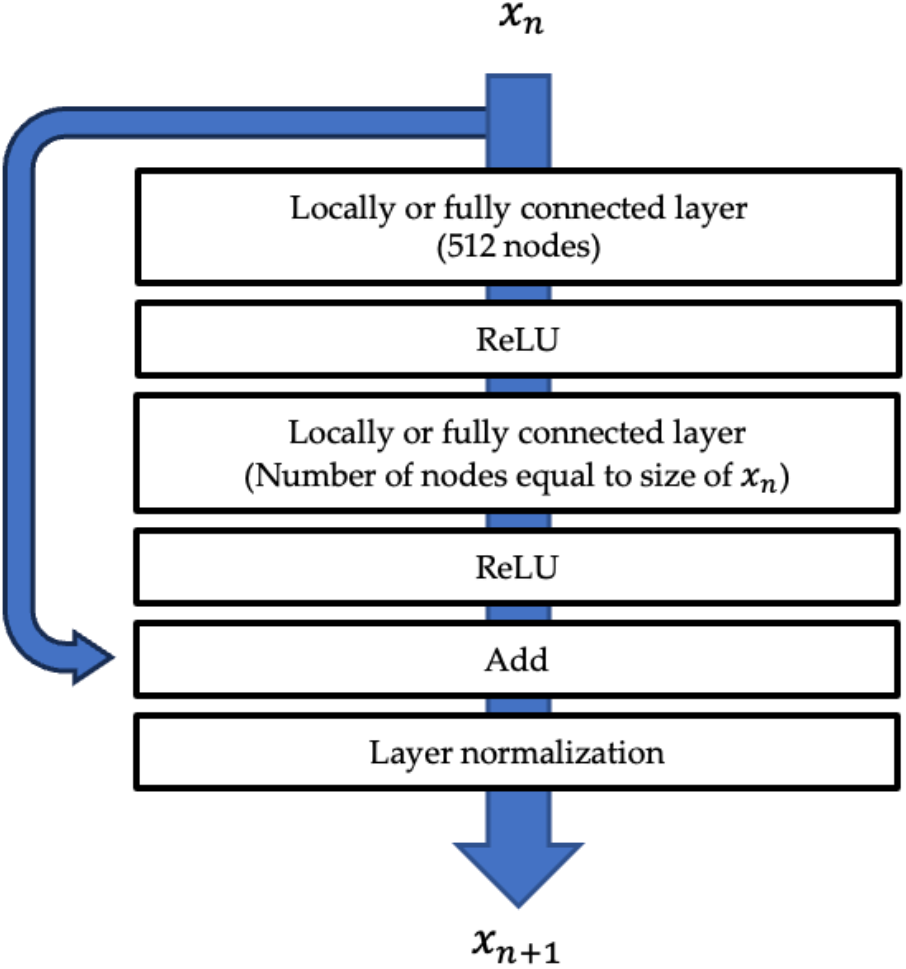
Illustration of a residual block. Depending on position in the encoder, there were locally or fully connected layers. In part two of the encoder, fully connected layers were applied channel-wise, while they were applied across all channels in part three and four. The first connected layer of the residual block always had 512 nodes. The second connected layer restored the data size to its input size to the block. ReLU activations were used after each connected layer. The original input ***x***_***n***_ was then added to the result of the connected layers and this was followed by layer normalization to produce the output ***x***_***n+*1**_ of the block.

In part two, channel-wise fully connected means that the same fully connected layer was applied to each channel, while locally connected means that each channel had its own connected layer. In parts three and four, the channels were analyzed together in fully connected layers. In part five, each cluster was analyzed separately using locally connected layers, except in the last layer, which was fully connected and produced the LDR.

To be able to extract the HDRs *Z* and *C*, the full encoder was implanted in three sections:1) part one, 2) part two, three and four, and 3) part five.

The intention with the architecture of the full encoder was that part one and two break down the signal into characteristic features for each channel. Based on these, part three classified the signal into one cluster, while part four analyzed and conveyed information of the mapping into lower dimensions. In part five, the outputs from part three and four were multiplied. Since part three only can produce one non-zero value, this selected information from part four and guaranteed the cluster encoding of part three was perpetuated.

There were two main problems arising from this architecture. One was that it was not guaranteed that input data would produce a non-zero output. The other was that the encoder may not use all possible clusters. There were several components in the loss function to counteract this. During training the batch size was 500 and the encoder had 10 clusters. The loss function then consisted of six parts:

1. To promote the encoder to learn to distribute data uniformly across clusters, the sum across the batch for each cluster for the output from part three was set to match 500/10 using the sum of the absolute error. The main reason for doing this was to promote the usage of all clusters, but also to avoid non-zero outputs.
2. To promote values above the ReLU threshold and for them to be closer to one, the sum for each example for the output from part three was set to match one using the sum of the absolute error. The main reason for doing this was to avoid non-zero outputs.
3. To promote values closer to one, a one hot encoding was created from the output of part three, and the representation *C* was set to match this using sum of the absolute error. This had an important effect on the separation of clusters.
4. There were three instances of matching representations using the binary cross-entropy:
  a. The sigmoid distribution of *Z* and t-distribution of *C*.
  b. The sigmoid distribution of *Z* and t-distribution of *Y*
  c. The sigmoid distribution of *C* and t-distribution of *Y*.

These subfunctions were weighted 10, 1, 1, 1, 1, and 1, respectively, and then summed to a total loss. Weighting part 1 higher was necessary to promote usage of all clusters.

The sigmoid distribution based on the ranked distances *r*_*ij*_ between example *i* and *j* was computed according to

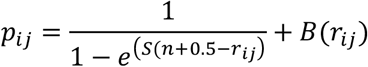

where *n* is the number of neighbors, the parameter *S* determine the slope, the term *B*(*r*_*ij*_) is used to provide the possibility to accentuate the difference between neighbors and non-neighbors with *B*(*r*_*ij*_) = *b, r*_*ih*_ ≤ *n* and *B*(*r*_*ij*_) = 0, *r*_*ih*_ > *n*, where *b* is a constant (examples of distributions using different parameter values are shown in Fig. 17).

**Fig. 17.**
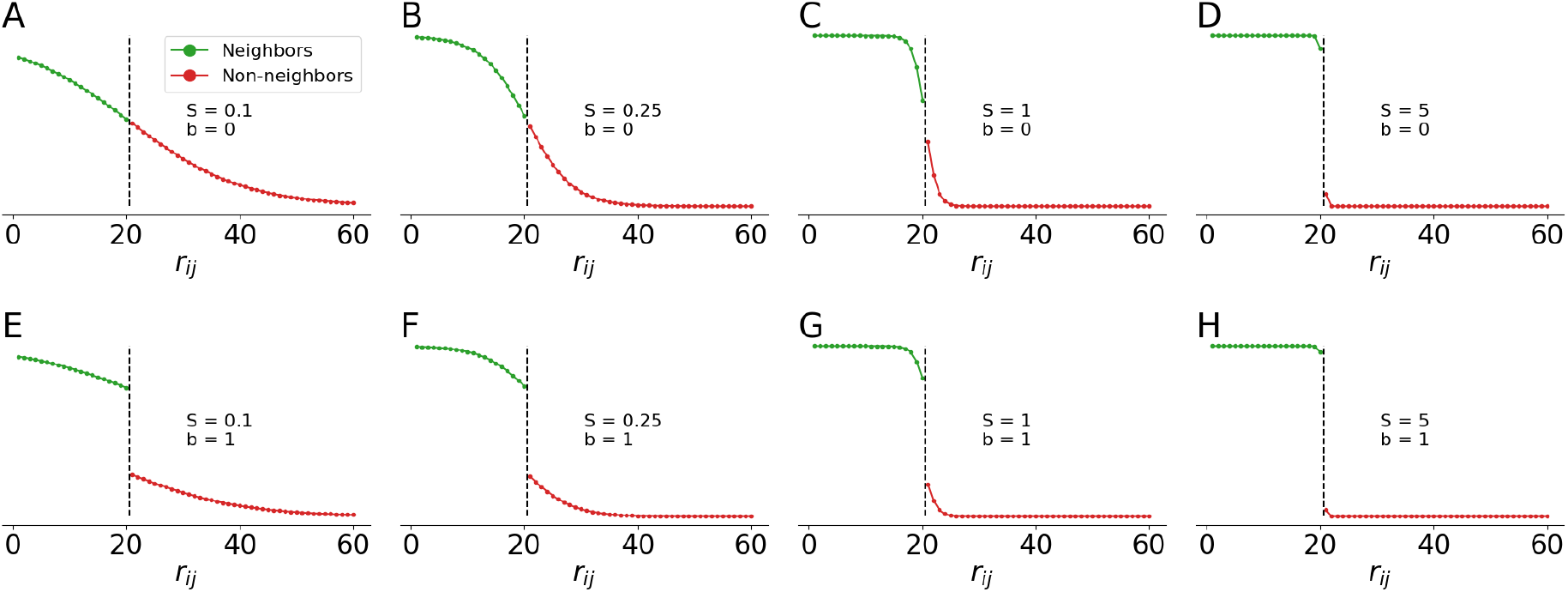
Illustration of the sigmoid distribution for 60 examples and 20 neighbors. Green: neighbors; red: non-neighbors, ***r***_***ij***_ is the ranked distances. (A–D) Effect of using ***S*** 0.1, 0.25, 1, or 5 and ***b*** = **0**. (E– H) Effect of using ***S*** 0.1, 0.25, 1, or 5 and ***b*** = **5**.

To further promote data points belonging to the same cluster being close, the ranked distances were conditional on cluster. This mean that during training, data points belonging to the same cluster were automatically ranked lower compared to the remaining data points. The distribution was computed before training and stored as an array, and during training examples could be assigned their values based on rank. Most of the processing time is due to computing the ranks, so using this approach does not add much time to training compared to using the two-valued distribution. In this project, parameters were set to *n* = 5, *S* = 1, and *b* = 1,000.

The encoder was trained on the PDs. Each example had a duration of 64 samples (256 ms). During training, the input EEG-interval to the encoder was randomly shifted ±16 samples. In parallel, the distance computed for the HDR was based on independently randomly shifted input EEG-interval. The intention was to dissociate the exact position of the marker from the HDR (Fig. 18).

**Fig. 18.**
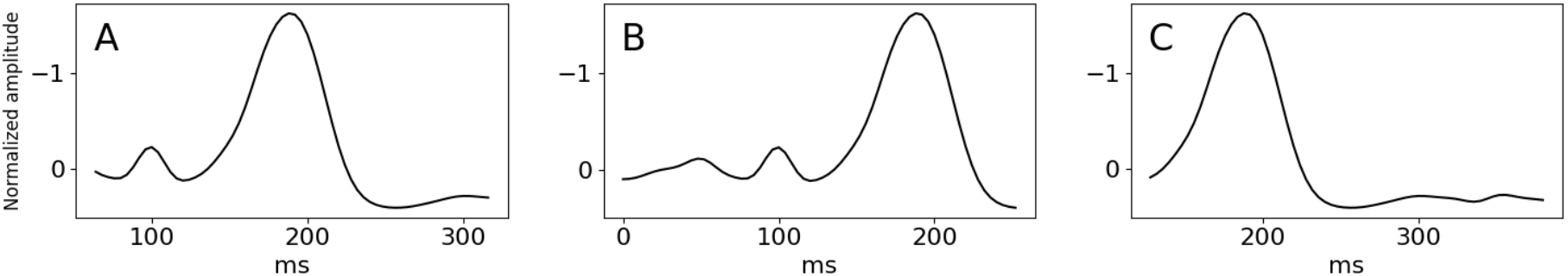
Example of the effect of shifting position of data. O1-electrode. (A) Waveform with centered position. (B) Waveform with maximum displacement backwards. (C) Waveform with maximum displacement forwards.

## Notes

### Competing Interest Statement

The authors have declared no competing interest.

### Funding Statement

Grants were received from Region Ostergotland (RO-974228, RO-962769, RO-941377, RO-986017, LIO-936176, and RO-941359). A.E. was supported by the ITEA3/VINNOVA funded project (AS-SIST).

## References

Abdalla M, Fine B. Hurdles to Artificial Intelligence Deployment: Noise in Schemas and “Gold” Labels. Radiol Artif Intell. 2023 Jan 11;5(2):e220056. doi: 10.1148/ryai.220056. PMID: 37035427; PMCID: PMC10077093.

Abend NS, Gutierrez-Colina A, Zhao H, Guo R, Marsh E, Clancy RR, Dlugos DJ. Interobserver reproducibility of electroencephalogram interpretation in critically ill children. J Clin Neurophysiol. 2011 Feb;28(1):15–9. doi: 10.1097/WNP.0b013e3182051123. PMID: 21221016; PMCID: PMC3107383.

Abend NS, Massey SL, Fitzgerald M, Fung F, Atkin NJ, Xiao R, Topjian AA. Interrater Agreement of EEG Interpretation After Pediatric Cardiac Arrest Using Standardized Critical Care EEG Terminology. J Clin Neurophysiology. 2017 Nov;34(6):534–541. doi: 10.1097/WNP.0000000000000424. PMID: 29023307; PMCID: PMC5679731.

Abou Jaoude M, Jing J, Sun H, Jacobs CS, Pellerin KR, Westover MB, Cash SS, Lam AD. Detection of mesial temporal lobe epileptiform discharges on intracranial electrodes using deep learning. Clin Neurophysiol. 2020 Jan;131(1):133–141. doi: 10.1016/j.clinph.2019.09.031. Epub 2019 Nov 11. PMID: 31760212; PMCID: PMC6879011.

Antoniades A, Spyrou L, Martin-Lopez D, Valentin A, Alarcon G, Sanei S, Cheong Took C. Detection of Interictal Discharges With Convolutional Neural Networks Using Discrete Ordered Multichannel Intracranial EEG. IEEE Trans Neural Syst Rehabil Eng. 2017 Dec;25(12):2285–2294. doi: 10.1109/TNSRE.2017.2755770. Epub 2017 Sep 22. PMID: 28952945.

Azuma H, Hori S, Nakanishi M, Fujimoto S, Ichikawa N, Furukawa TA. An intervention to improve the interrater reliability of clinical EEG interpretations. Psychiatry Clin Neurosci. 2003 Oct;57(5):485–9. doi: 10.1046/j.1440-1819.2003.01152.x. PMID: 12950702.

Bagheri E, Dauwels J, Dean BC, Waters CG, Westover MB, Halford JJ. Interictal epileptiform discharge characteristics underlying expert interrater agreement. Clin Neurophysiol. 2017 Oct;128(10):1994–2005. doi: 10.1016/j.clinph.2017.06.252. Epub 2017 Jul 18. PMID: 28837905; PMCID: PMC5842710.

Beuchat I, Alloussi S, Reif PS, Sterlepper N, Rosenow F, Strzelczyk A. Prospective evaluation of interrater agreement between EEG technologists and neurophysiologists. Sci Rep. 2021 Jun 28;11(1):13406. doi: 10.1038/s41598-021-92827-3. PMID: 34183718; PMCID: PMC8238944.

Black MA, Jones RD, Carroll GJ, Dingle AA, Donaldson IM, Parkin PJ. Real-time detection of epileptiform activity in the EEG: a blinded clinical trial. Clin Electroencephalogr. 2000 Jul;31(3):122–30. doi: 10.1177/155005940003100304. PMID: 10923198.

Chen L, Ho CK, Lam VK, Fong SY, Li AM, Lam SP, Wing YK. Interrater and intrarater reliability in multiple sleep latency test. J Clin Neurophysiol. 2008 Aug;25(4):218–21. doi: 10.1097/WNP.0b013e31817f36a6. PMID: 18677186.

Cheng C, Liu Y, You B, Zhou Y, Gao F, Yang L, Dai Y. Multilevel Feature Learning Method for Accurate Interictal Epileptiform Spike Detection. IEEE Trans Neural Syst Rehabil Eng. 2022;30:2506– 2516. doi: 10.1109/TNSRE.2022.3193666. Epub 2022 Sep 8. PMID: 35877795.

Cohen JA. Coefficient of Agreement for Nominal Scales. Educational and Psychological Measurement. 1960, 20(1), 37–46. 10.1177/001316446002000104

Chung YG, Lee WJ, Na SM, Kim H, Hwang H, Yun CH, Kim KJ. Deep learning-based automated detection and multiclass classification of focal interictal epileptiform discharges in scalp electroencephalograms. Sci Rep. 2023 Apr 25;13(1):6755. doi: 10.1038/s41598-023-33906-5. PMID: 37185941; PMCID: PMC10130023.

Craik A, He Y, Contreras-Vidal JL. Deep learning for electroencephalogram (EEG) classification tasks: a review. J Neural Eng. 2019 Jun;16(3):031001. doi: 10.1088/1741-2552/ab0ab5. Epub 2019 Feb 26. PMID: 30808014.

Dümpelmann M, Elger CE. Visual and automatic investigation of epileptiform spikes in intracranial EEG recordings. Epilepsia. 1999 Mar;40(3):275–85. doi: 10.1111/j.1528-1157.1999.tb00704.x. PMID: 10080505.

Faghihpirayesh R, Ruf S, Rocca M, Garner R, Vespa P, Erdogmus D, Duncan D. Automatic Detection of EEG Epileptiform Abnormalities in Traumatic Brain Injury using Deep Learning. Annu Int Conf IEEE Eng Med Biol Soc. 2021 Nov;2021:302–305. doi: 10.1109/EMBC46164.2021.9630242. PMID: 34891296; PMCID: PMC8860400.

Fleiss JL. Measuring nominal scale agreement among many raters. Psychological Bulletin. 1971, 76(5), 378–382. 10.1037/h0031619.

Fürbass F, Kural MA, Gritsch G, Hartmann M, Kluge T, Beniczky S. An artificial intelligence-based EEG algorithm for detection of epileptiform EEG discharges: Validation against the diagnostic gold standard. Clin Neurophysiol. 2020 Jun;131(6):1174–1179. doi: 10.1016/j.clinph.2020.02.032. Epub 2020 Apr 2. PMID: 32299000.

Gaspard N, Hirsch LJ, LaRoche SM, Hahn CD, Westover MB; Critical Care EEG Monitoring Research Consortium. Interrater agreement for Critical Care EEG Terminology. Epilepsia. 2014 Sep;55(9):1366–73. doi: 10.1111/epi.12653. Epub 2014 Jun 2. PMID: 24888711; PMCID: PMC4879939.

Geng D, Alkhachroum A, Melo Bicchi MA, Jagid JR, Cajigas I, Chen ZS. Deep learning for robust detection of interictal epileptiform discharges. J Neural Eng. 2021 Apr 8;18(5). doi: 10.1088/1741-2552/abf28e. PMID: 33770777.

Gerber PA, Chapman KE, Chung SS, Drees C, Maganti RK, Ng YT, Treiman DM, Little AS, Kerrigan JF. Interobserver agreement in the interpretation of EEG patterns in critically ill adults. J Clin Neurophysiol. 2008 Oct;25(5):241–9. doi: 10.1097/WNP.0b013e318182ed67. PMID: 18791475.

Gwet KL. Computing inter-rater reliability and its variance in the presence of high agreement. British Journal of Mathematical and Statistical Psychology. 2008, 61: 29–48. 10.1348/000711006X126600

Gwet KL. Handbook of Interrater Reliability. 2014, 4th edition, pp 57–58, Advanced Analytics, LLC.

Halford JJ. Computerized epileptiform transient detection in the scalp electroencephalogram: obstacles to progress and the example of computerized ECG interpretation. Clin Neurophysiol. 2009 Nov;120(11):1909–1915. doi: 10.1016/j.clinph.2009.08.007. Epub 2009 Oct 15. PMID: 19836303.

Halford JJ, Schalkoff RJ, Zhou J, Benbadis SR, Tatum WO, Turner RP, Sinha SR, Fountain NB, Arain A, Pritchard PB, Kutluay E, Martz G, Edwards JC, Waters C, Dean BC. Standardized database development for EEG epileptiform transient detection: EEGnet scoring system and machine learning analysis. J Neurosci Methods. 2013 Jan 30;212(2):308–16. doi: 10.1016/j.jneumeth.2012.11.005. Epub 2012 Nov 19. PMID: 23174094.

Halford JJ, Arain A, Kalamangalam GP, LaRoche SM, Leonardo B, Basha M, Azar NJ, Kutluay E, Martz GU, Bethany WJ, Waters CG, Dean BC. Characteristics of EEG Interpreters Associated With Higher Interrater Agreement. J Clin Neurophysiol. 2017 Mar;34(2):168–173. doi: 10.1097/WNP.0000000000000344. PMID: 27662336; PMCID: PMC5784780.

Halford JJ, Westover MB, LaRoche SM, Macken MP, Kutluay E, Edwards JC, Bonilha L, Kalamangalam GP, Ding K, Hopp JL, Arain A, Dawson RA, Martz GU, Wolf BJ, Waters CG, Dean BC. Interictal Epileptiform Discharge Detection in EEG in Different Practice Settings. J Clin Neurophysiol. 2018 Sep;35(5):375–380. doi: 10.1097/WNP.0000000000000492. PMID: 30028830; PMCID: PMC6126936.

Hussain SA, Kwong G, Millichap JJ, Mytinger JR, Ryan N, Matsumoto JH, Wu JY, Lerner JT, Sankar R. Hypsarrhythmia assessment exhibits poor interrater reliability: a threat to clinical trial validity. Epilepsia. 2015 Jan;56(1):77–81. doi: 10.1111/epi.12861. Epub 2014 Nov 10. PMID: 25385396.

Jeon Y, Chung YG, Joo T, Kim H, Hwang H, Kim KJ. Deep Learning-Based Detection of Epileptiform Discharges for Self-Limited Epilepsy With Centrotemporal Spikes. IEEE Trans Neural Syst Rehabil Eng. 2022;30:2939–2949. doi: 10.1109/TNSRE.2022.3215526. Epub 2022 Oct 27. PMID: 36260578.

Jiang L, Fan Q, Ren J, Dong F, Jiang T, Liu J. An improved BECT spike detection method with functional brain network features based on PLV. Front Neurosci. 2023 Mar 16;17:1150668. doi: 10.3389/fnins.2023.1150668. PMID: 37008227; PMCID: PMC10060895.

Jing J, Herlopian A, Karakis I, Ng M, Halford JJ, Lam A, Maus D, Chan F, Dolatshahi M, Muniz CF, Chu C, Sacca V, Pathmanathan J, Ge W, Sun H, Dauwels J, Cole AJ, Hoch DB, Cash SS, Westover MB. Interrater Reliability of Experts in Identifying Interictal Epileptiform Discharges in Electroencephalograms. JAMA Neurol. 2020 Jan 1;77(1):49–57. doi: 10.1001/jamaneurol.2019.3531. Erratum in: JAMA Neurol. 2020 Jan 1;77(1):135. PMID: 31633742; PMCID: PMC6806666.

Johansen AR, Jin J, Maszczyk T, Dauwels J, Cash SS, Westover MB. EPILEPTIFORM SPIKE DETECTION VIA CONVOLUTIONAL NEURAL NETWORKS. Proc IEEE Int Conf Acoust Speech Signal Process. 2016 Mar;2016:754–758. doi: 10.1109/ICASSP.2016.7471776. Epub 2016 May 19. PMID: 29527131; PMCID: PMC5842703.

Joshi CN, Chapman KE, Bear JJ, Wilson SB, Walleigh DJ, Scheuer ML. Semiautomated Spike Detection Software Persyst 13 Is Noninferior to Human Readers When Calculating the Spike-Wave Index in Electrical Status Epilepticus in Sleep. J Clin Neurophysiol. 2018 Sep;35(5):370–374. doi: 10.1097/WNP.0000000000000493. PMID: 29933261.

Karimi D, Dou H, Warfield SK, Gholipour A. Deep learning with noisy labels: Exploring techniques and remedies in medical image analysis. Med Image Anal. 2020 Oct;65:101759. doi: 10.1016/j.media.2020.101759. Epub 2020 Jun 20. PMID: 32623277; PMCID: PMC7484266.

Kingma DP, Ba J. Adam: A Method for Stochastic Optimization. In Proceedings of the International Conference on Learn-ing Representations, San Diego, CA, USA, 7–9 May 2015.

Klambauer G, Unterthiner T, Mayr A, Hochreiter S. Self-normalizing neural networks. In Proceedings of the 31st International Conference on Neural Information Processing Systems (NIPS’17). 2017. Curran Associates Inc., Red Hook, NY, USA, 972–981.

Kural MA, Jing J, Fürbass F, Perko H, Qerama E, Johnsen B, Fuchs S, Westover MB, Beniczky S. Accurate identification of EEG recordings with interictal epileptiform discharges using a hybrid approach: Artificial intelligence supervised by human experts. Epilepsia. 2022 May;63(5):1064–1073. doi: 10.1111/epi.17206. Epub 2022 Mar 7. PMID: 35184276; PMCID: PMC9148170.

Landis JR, Koch GG. The measurement of observer agreement for categorical data. Biometrics. 1977 Mar;33(1):159–74. PMID: 843571.

Mani R, Arif H, Hirsch LJ, Gerard EE, LaRoche SM. Interrater reliability of ICU EEG research terminology. J Clin Neurophysiol. 2012 Jun;29(3):203–12. doi: 10.1097/WNP.0b013e3182570f83. PMID: 22659712.

Massey SL, Shou H, Clancy R, DiGiovine M, Fitzgerald MP, Fung FW, Farrar J, Abend NS. Interrater and Intrarater Agreement in Neonatal Electroencephalogram Background Scoring. J Clin Neurophysiol. 2019 Jan;36(1):1–8. doi: 10.1097/WNP.0000000000000534. PMID: 30383719; PMCID: PMC6322680.

McDougall M, Albaqami H, Mubashar Hassan G, Datta A. Patient Independent Interictal Epileptiform Discharge Detection. Annu Int Conf IEEE Eng Med Biol Soc. 2023 Jul;2023:1–6. doi: 10.1109/EMBC40787.2023.10341194. PMID: 38082573.

Medvedev AV, Agoureeva GI, Murro AM. A Long Short-Term Memory neural network for the detection of epileptiform spikes and high frequency oscillations. Sci Rep. 2019 Dec 18;9(1):19374. doi: 10.1038/s41598-019-55861-w. PMID: 31852929; PMCID: PMC6920137.

Mytinger JR, Weber A, Vidaurre J. High Amplitude Background Slow Waves in Normal Children Aged 3 to 18 Months: Implications for the Consideration of Hypsarhythmia. J Clin Neurophysiol. 2018 Mar;35(2):151–154. doi: 10.1097/WNP.0000000000000449. PMID: 29315089.

Nejedly P, Kremen V, Lepkova K, Mivalt F, Sladky V, Pridalova T, Plesinger F, Jurak P, Pail M, Brazdil M, Klimes P, Worrell G. Utilization of temporal autoencoder for semi-supervised intracranial EEG clustering and classification. Sci Rep. 2023 Jan 13;13(1):744. doi: 10.1038/s41598-023-27978-6. PMID: 36639549; PMCID: PMC9839708.

Nguyen DK, Girard ME, Cossette P, Saint-Hilaire JM. Audit of EEG reporting temporal abnormalities. Can J Neurol Sci. 2010 Nov;37(6):819–25. doi: 10.1017/s0317167100051507. PMID: 21059545.

Nizam A, Chen S, Wong S. Best-case kappa scores calculated retrospectively from EEG report databases. J Clin Neurophysiol. 2013 Jun;30(3):268–74. doi: 10.1097/WNP.0b013e3182933da7. PMID: 23733091.

Nordli DR Jr, Bazil CW, Scheuer ML, Pedley TA. Recognition and classification of seizures in infants. Epilepsia. 1997 May;38(5):553–60. doi: 10.1111/j.1528-1157.1997.tb01140.x. PMID: 9184601.

Nhu D, Janmohamed M, Antonic-Baker A, Perucca P, O’Brien TJ, Gilligan AK, Kwan P, Tan CW, Kuhlmann L. Deep learning for automated epileptiform discharge detection from scalp EEG: A systematic review. J Neural Eng. 2022 Oct 19;19(5). doi: 10.1088/1741-2552/ac9644. PMID: 36174541.

Nhu D, Janmohamed M, Shakhatreh L, Gonen O, Perucca P, Gilligan A, Kwan P, O’Brien TJ, Tan CW, Kuhlmann L. Automated Interictal Epileptiform Discharge Detection from Scalp EEG Using Scalable Time-series Classification Approaches. Int J Neural Syst. 2023 Jan;33(1):2350001. doi: 10.1142/S0129065723500016. Epub 2023 Jan 5. PMID: 36599664.

Obeid I, Picone J. The Temple University Hospital EEG Data Corpus. Front Neurosci. 2016 May 13;10:196. doi: 10.3389/fnins.2016.00196. PMID: 27242402; PMCID: PMC4865520.

Pedregosa F, Varoquaux G, Gramfort A, Michel V, Thirion B, Grisel O, Blondel M, Prettenhofer P, Weiss R, Dubourg V, Vanderplas J, Passos A, Cournapeau D, Brucher M, Perrot M, Duchesnay E. Scikitlearn: Machine learning in Python. Journal of Machine Learning Research. 2011, 12(Oct), 2825–2830.

Piccinelli P, Viri M, Zucca C, Borgatti R, Romeo A, Giordano L, Balottin U, Beghi E. Inter-rater reliability of the EEG reading in patients with childhood idiopathic epilepsy. Epilepsy Res. 2005 Aug-Sep;66(1-3):195–8. doi: 10.1016/j.eplepsyres.2005.07.004. PMID: 16118044.

Reus EEM, Visser GH, Cox FME. Using sampled visual EEG review in combination with automated detection software at the EMU. Seizure. 2020 Aug;80:96–99. doi: 10.1016/j.seizure.2020.06.002. Epub 2020 Jun 4. PMID: 32554293.

Reus EEM, Visser GH, Cox FME. Determining the Spike-Wave Index Using Automated Detection Software. J Clin Neurophysiol. 2021 May 1;38(3):198–201. doi: 10.1097/WNP.0000000000000672. PMID: 31834040.

Reus EEM, Cox FME, van Dijk JG, Visser GH. Automated spike detection: Which software package? Seizure. 2022 Feb;95:33–37. doi: 10.1016/j.seizure.2021.12.012. Epub 2021 Dec 26. PMID: 34974231.

Ronner HE, Ponten SC, Stam CJ, Uitdehaag BM. Inter-observer variability of the EEG diagnosis of seizures in comatose patients. Seizure. 2009 May;18(4):257–63. doi: 10.1016/j.seizure.2008.10.010. Epub 2008 Nov 28. PMID: 19046902.

Roy Y, Banville H, Albuquerque I, Gramfort A, Falk TH, Faubert J. Deep learning-based electro-encephalography analysis: a systematic review. J Neural Eng. 2019 Aug 14;16(5):051001. doi: 10.1088/1741-2552/ab260c. PMID: 31151119.

Stroink H, Schimsheimer RJ, de Weerd AW, Geerts AT, Arts WF, Peeters EA, Brouwer OF, Boudewijn Peters A, van Donselaar CA. Interobserver reliability of visual interpretation of electroencephalograms in children with newly diagnosed seizures. Dev Med Child Neurol. 2006 May;48(5):374–7. doi: 10.1017/S0012162206000806. PMID: 16608546.

Svantesson M, Olausson H, Eklund A, Thordstein M. Get a New Perspective on EEG: Convolutional Neural Network Encoders for Parametric t-SNE. Brain Sci. 2023 Mar 7;13(3):453. doi: 10.3390/brainsci13030453. PMID: 36979263; PMCID: PMC10046040.

Svantesson M, Olausson H, Eklund A, Thordstein M. Virtual EEG-electrodes: Convolutional neural networks as a method for upsampling or restoring channels. J Neurosci Methods. 2021 May 1;355:109126. doi: 10.1016/j.jneumeth.2021.109126. Epub 2021 Mar 9. PMID: 33711358.

Thangavel P, Thomas J, Peh WY, Jing J, Yuvaraj R, Cash SS, Chaudhari R, Karia S, Rathakrishnan R, Saini V, Shah N, Srivastava R, Tan YL, Westover B, Dauwels J. Time-Frequency Decomposition of Scalp Electroencephalograms Improves Deep Learning-Based Epilepsy Diagnosis. Int J Neural Syst. 2021 Aug;31(8):2150032. doi: 10.1142/S0129065721500325. Epub 2021 Jul 16. PMID: 34278972; PMCID: PMC9340811.

Tjepkema-Cloostermans MC, de Carvalho RCV, van Putten MJAM. Deep learning for detection of focal epileptiform discharges from scalp EEG recordings. Clin Neurophysiol. 2018 Oct;129(10):2191– 2196. doi: 10.1016/j.clinph.2018.06.024. Epub 2018 Jul 9. PMID: 30025804.

Thomas J, Jin J, Thangavel P, Bagheri E, Yuvaraj R, Dauwels J, Rathakrishnan R, Halford JJ, Cash SS, Westover B. Automated Detection of Interictal Epileptiform Discharges from Scalp Electroencephalograms by Convolutional Neural Networks. Int J Neural Syst. 2020 Nov;30(11):2050030. doi: 10.1142/S0129065720500306. Epub 2020 Aug 19. PMID: 32812468; PMCID: PMC7606586.

Thomas J, Thangavel P, Peh WY, Jing J, Yuvaraj R, Cash SS, Chaudhari R, Karia S, Rathakrishnan R, Saini V, Shah N, Srivastava R, Tan YL, Westover B, Dauwels J. Automated Adult Epilepsy Diagnostic Tool Based on Interictal Scalp Electroencephalogram Characteristics: A Six-Center Study. Int J Neural Syst. 2021 May;31(5):2050074. doi: 10.1142/S0129065720500744. Epub 2021 Jan 12. PMID: 33438530; PMCID: PMC9343226.

Vach W, Gerke O. Gwet’s AC1 is not a substitute for Cohen’s kappa - A comparison of basic properties. MethodsX. 2023 May 10;10:102212. doi: 10.1016/j.mex.2023.102212. PMID: 37234937; PMCID: PMC10205778.

van der Maaten L, Hinton G. Visualizing Data using t-SNE. Journal of Machine Learning Research. 2008, 9, 2579--2605.

van der Maaten L. Learning a Parametric Embedding by Preserving Local Structure. Proceedings of the Twelfth International Conference on Artificial Intelligence and Statistics. Proceedings of Machine Learning Research. 2009, 5:384–391.

Vaswani A, Shazeer N, Parmar N, Uszkoreit J, Jones L, Gomez AN, Kaiser Ł, Polosukhin I. Attention is all you need. In Proceedings of the 31st International Conference on Neural Information Processing Systems (NIPS’17). 2017. Curran Associates Inc., Red Hook, NY, USA, 6000–6010.

Virtanen P, Gommers R, Oliphant TE, Haberland M, Reddy T, Cournapeau D, Burovski E, Peterson P, Weckesser W, Bright J, et al. SciPy 1.0: Fundamental Algorithms for Scientific Computing in Python. Nature Methods. 2020 17(3), 261–272.

Wang X, Wang X, Wang C, Wang Z, Liu X, Lv X, Tang Y. A Two-Stage Automatic System for Detection of Interictal Epileptiform Discharges from Scalp Electroencephalograms. eNeuro. 2023 Nov 21;10(11):ENEURO.0111-23.2023. doi: 10.1523/ENEURO.0111-23.2023. PMID: 37914407; PMCID: PMC10668214.

Wei B, Zhao X, Shi L, Xu L, Liu T, Zhang J. A deep learning framework with multi-perspective fusion for interictal epileptiform discharges detection in scalp electroencephalogram. J Neural Eng. 2021 Jul 21;18(4). doi: 10.1088/1741-2552/ac0d60. PMID: 34157696.

Wilson SB, Harner RN, Duffy FH, Tharp BR, Nuwer MR, Sperling MR. Spike detection. I. Correlation and reliability of human experts. Electroencephalogr Clin Neurophysiol. 1996 Mar;98(3):186–98. doi: 10.1016/0013-4694(95)00221-9. PMID: 8631278.

Young GB, McLachlan RS, Kreeft JH, Demelo JD. An electroencephalographic classification for coma. Can J Neurol Sci. 1997 Nov;24(4):320–5. doi: 10.1017/s0317167100032996. PMID: 9398979.

Zhang L, Wang X, Jiang J, Xiao N, Guo J, Zhuang K, Li L, Yu H, Wu T, Zheng M, Chen D. Automatic interictal epileptiform discharge (IED) detection based on convolutional neural network (CNN). Front Mol Biosci. 2023 Apr 7;10:1146606. doi: 10.3389/fmolb.2023.1146606. PMID: 37091867; PMCID: PMC10119410.

Zhuo Ding J, Mallick R, Carpentier J, McBain K, Gaspard N, Brandon Westover M, Fantaneanu TA. Resident training and interrater agreements using the ACNS critical care EEG terminology. Seizure. 2019 Mar;66:76–80. doi: 10.1016/j.seizure.2019.02.013. Epub 2019 Feb 20. PMID: 30818180; PMCID: PMC6778405.

